# Distinguishing features of Long COVID identified through immune profiling

**DOI:** 10.1101/2022.08.09.22278592

**Authors:** Jon Klein, Jamie Wood, Jillian Jaycox, Peiwen Lu, Rahul M. Dhodapkar, Jeff R. Gehlhausen, Alexandra Tabachnikova, Laura Tabacof, Amyn A. Malik, Kathy Kamath, Kerrie Greene, Valter Silva Monteiro, Mario Peña-Hernandez, Tianyang Mao, Bornali Bhattacharjee, Takehiro Takahashi, Carolina Lucas, Julio Silva, Dayna Mccarthy, Erica Breyman, Jenna Tosto-Mancuso, Yile Dai, Emily Perotti, Koray Akduman, Tiffany J. Tzeng, Lan Xu, Inci Yildirim, Harlan M. Krumholz, John Shon, Ruslan Medzhitov, Saad B. Omer, David van Dijk, Aaron M. Ring, David Putrino, Akiko Iwasaki

## Abstract

SARS-CoV-2 infection can result in the development of a constellation of persistent sequelae following acute disease called post-acute sequelae of COVID-19 (PASC) or Long COVID^1–3^. Individuals diagnosed with Long COVID frequently report unremitting fatigue, post-exertional malaise, and a variety of cognitive and autonomic dysfunctions^1–3^; however, the basic biological mechanisms responsible for these debilitating symptoms are unclear. Here, 215 individuals were included in an exploratory, cross-sectional study to perform multi-dimensional immune phenotyping in conjunction with machine learning methods to identify key immunological features distinguishing Long COVID. Marked differences were noted in specific circulating myeloid and lymphocyte populations relative to matched control groups, as well as evidence of elevated humoral responses directed against SARS-CoV-2 among participants with Long COVID. Further, unexpected increases were observed in antibody responses directed against non-SARS-CoV-2 viral pathogens, particularly Epstein-Barr virus. Analysis of circulating immune mediators and various hormones also revealed pronounced differences, with levels of cortisol being uniformly lower among participants with Long COVID relative to matched control groups. Integration of immune phenotyping data into unbiased machine learning models identified significant distinguishing features critical in accurate classification of Long COVID, with decreased levels of cortisol being the most significant individual predictor. These findings will help guide additional studies into the pathobiology of Long COVID and may aid in the future development of objective biomarkers for Long COVID.

## Introduction

Recovery from viral infections is heterogeneous and chronic symptoms may persist in a subset of convalescent individuals. Clinical sequelae can manifest following a variety of acute infections across a diverse range of viral families^4–13^. Moreover, post-acute infection syndromes (PAIS) following acute viral diseases have been described for more than a century^14–17^. Yet despite their ubiquity and historical record, the basic biology underlying the development of PAIS following viral infections remains unclear^18^.

SARS-CoV-2 is a zoonotic betacoronavirus responsible for more than 6 million deaths since its initial detection in late 2019^19^. The acute phase of COVID-19 has been studied extensively and in severe cases presents with extensive immunological and multi-organ system dysfunction^20–24^. Outcomes following COVID-19 are varied, ranging from complete recovery to a significantly increased risk of an assortment of adverse clinical events - even among those with initially mild disease^2, 25^. Subsets of convalescent COVID-19 patients may also develop new or aggravated sequelae for months to years following resolution of acute COVID-19, comprising a nascent clinical syndrome known as post-acute sequelae of COVID-19 (PASC) or Long COVID. Clinically, Long COVID presents as a constellation of debilitating symptoms most commonly including unremitting fatigue, post-exertional malaise, cognitive impairment, and autonomic dysfunction among many others^1, 3, 26–29^. Estimates of the prevalence of Long COVID vary substantially^30, 31^; however, even the most optimistic estimates of prevalence present an enormous burden on millions of people with significant clinical, social, and economic impacts given the global breadth of SARS-CoV-2 exposure. While the underlying pathogenesis of Long COVID remains unclear, current hypotheses include persistent virus or viral remnants, autoimmunity, dysbiosis, latent virus reactivation, and tissue damage caused by lingering inflammation^18, 32–38^.

To interrogate the biological underpinnings of Long COVID, an exploratory cross-sectional study was designed (Mount Sinai-Yale Long COVID or ‘MY-LC’) involving 215 participants composed of four groups: (1) healthy, uninfected controls (Healthy Controls or ‘HC’); (2) healthy, unvaccinated, previously SARS-CoV-2-infected historical controls (Healthcare Workers or ‘HCW’); (3) healthy, previously SARS-CoV-2 infected controls without persistent symptoms (Convalescent Controls or ‘CC’); and (4) individuals with persistent symptoms following acute SARS-CoV-2 infection (Long COVID or ‘LC’). Among the HCW, CC, and LC groups, enrolled participants were primarily non-hospitalized during acute COVID-19, and CC and LC groups were on average more than one year from the initial infection. From each group, systematic, multi-dimensional immunophenotyping and unbiased machine learning of aggregated data was performed to identify potential biomarkers of Long COVID.

## Results

### Clinical characterization and demographic analysis of MY-LC groups

The MY-LC Study enrolled a total of 220 participants (101 LC, 41 CC, 41 HC, and 37 HCW) at two study sites (HC, CC, and LC from Mount Sinai Hospital, New York City, New York, and HCW from Yale New Haven Hospital, New Haven, Connecticut). After initial enrollment and preliminary review of electronic medical records, two participants were excluded from the Long COVID group (n = 2 [1.98%] for pharmacologic immunosuppression secondary to primary immune deficiency and solid organ transplant), one from the healthy group (n = 1 [2.38%] for current pregnancy), and two from the convalescent group (n = 2 [4.76%] for current pregnancy and monogenic disorder). After exclusion, the final group sizes were 99 individuals in LC, 40 in HC, 39 in CC, and 37 in HCW for a total study size of 215 individuals. (Fig. 1A). There were no significant differences in the proportion of excluded participants between groups (p = .6255 [Chi-square **χ**: 0.9386, d.f. = 2]).

**Figure 1.**
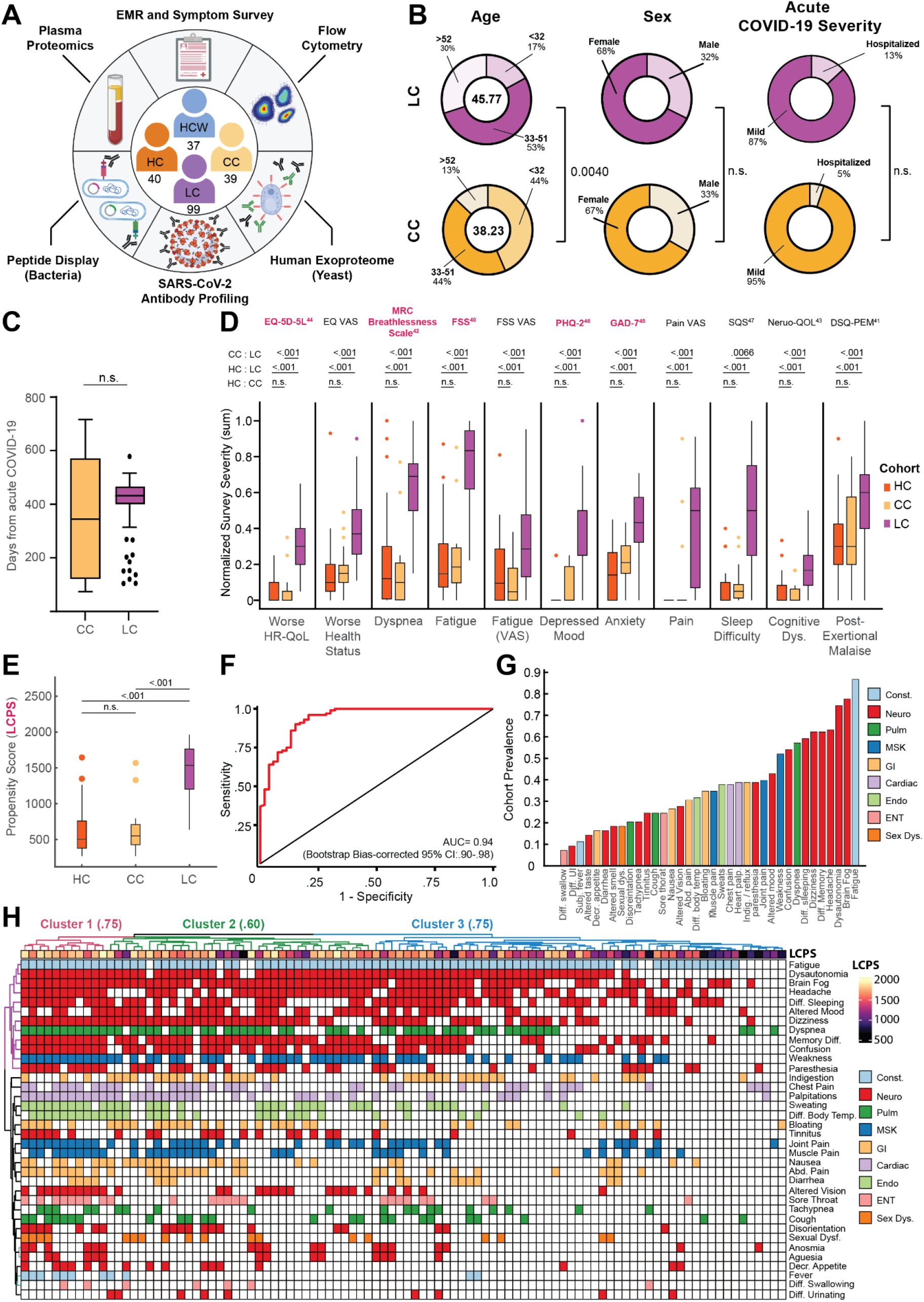
Demographic and clinical stratification of participants with Long COVID. (**A**) Schematic of MY-LC study design. Numbers in center of diagram indicate enrolled participants in each study group (*clockwise,* ‘HCW’ = historical, unvaccinated SARS-CoV-2 infected controls, ‘CC’ = convalescent SARS-CoV-2 individuals without persistent symptoms; ‘LC’ = convalescent SARS-CoV-2 individuals with persistent symptoms; ‘HC’ = healthy controls with no prior-SARS-CoV-2 infection). Outer ring describes different assay modalities performed on samples from each group. (**B**) Demographic features of interest for LC (top row, purple) and CC (bottom row, yellow) displayed as ring charts. Center values in ‘Age’ column are group average ages in years. Statistical significance is reported by brackets as relevant post-hoc comparisons (‘Age’) or Chi-square tests (‘Sex’ and ‘Acute Disease Severity’) and are further detailed in Extended Table 2. **(C)** Box plots of days from acute symptom onset between CC and LC groups. Central lines indicate group medians, with top and bottom lines indicating 75^th^ and 25^th^ percentiles, respectively. Significance was assessed using Wilcoxon Rank-Sum testing. **(D)** Box plots of Min-Max, oriented and normalized survey responses for HC (orange), CC (yellow), and LC (purple) groups. Individual survey instruments are arranged in columns, with formal names displayed in bold text (top) and respective health dimensions reported along corresponding points of x-axis. Surveys marked in red were combined to generate Long COVID Propensity Scores (LCPS). Central lines of box plots indicate group medians, with top and bottom lines indicating 75^th^ and 25^th^ percentiles, respectively. Significance was assessed using Kruskal-Wallis tests with post-hoc comparisons using Dunn’s test reported along upper rows (top to bottom: (1) CC vs. LC (2) HC vs. LC (3) HC vs. CC) and are corrected for multiple comparisons using Tukey’s Method. Superscripts refer to ‘Reference’ entries for previously published, validated survey instruments. **(E)** Box plots of Long COVID Propensity Scores (LCPS). Central lines indicate group medians, with top and bottom lines indicating 75^th^ and 25^th^ percentiles, respectively. Significance was assessed using Kruskal-Wallis tests with post-hoc comparisons using Dunn’s test and are corrected for multiple comparisons using Tukey’s Method. **(F)** Receiver-Operator Curve (ROC) analysis of LCPS scores. Area under the curve (AUC) is reported with Bootstrap Bias-corrected 95% confidence intervals of AUC. **(G)** Prevalence of top 30 self-reported binary symptoms ranked from most prevalent (right) to least prevalent (left). Symptoms are colored according to investigator-assigned physiological systems: ‘Const. / Constitutional’ = light blue, ‘Neuro. / Neurological’ = red, ‘Pulm. / Pulmonary’ = green, ‘MSK / Musculoskeletal’ = blue, ‘GI / Gastrointestinal’ = tan, ‘Cardiac’ = light purple, ‘Endo. / Endocrine’ = light green, ‘ENT / Ear, Nose, Throat’ = pink, and ‘Sex Dys. / Sexual Dysfunction’ = orange. **(H)** Heatmap of self-reported binary symptoms bi-clustered by Hamming distances (rows and columns) and colored according to physiological system as previous (G, before). Columns are annotated by LCPS scores with bootstrapped cluster reproducibility scores reported in parentheses (bootstrapped Jaccard similarity). For all panels, only significant differences between groups are displayed. *Abbreviations: EMR = electronic medical record; n.s. = not significant; Dys. = dysfunction; CI = confidence interval; Diff. = difficulty; UI = urination; Subj. = subjective; Decr. = decreased; Abd. = abdominal; Indig. = indigestion*.

Initial comparison of demographic factors demonstrated that the Long COVID and Convalescent groups were well-matched in age (mean 45.77 years old LC vs. mean 38.23 years old CC; Kruskal-Wallis post-hoc p = 0.004), sex (68% female LC vs. 67% female CC, p = .9465 [**χ**: 0.1101, d.f. = 2]), and proportion of hospitalized acute COVID-19 (13.13% LC vs. 5.13% CC; Fisher’s Exact p = .2324 [OR: 2.797, 95% CI 0.66 – 12.90]) (Fig. 1B). For Long COVID and Convalescent groups, analysis of elapsed days since initial SARS-CoV-2 infection revealed no significant difference in median times from acute disease (432 days LC vs. 344 days CC; Mann-Whitney p = .0572) (Fig. 1C), further enabling direct comparison of persistent symptom and convalescent groups. Additionally, the majority of acute SARS-CoV-2 infections within the Long COVID group (76%) occurred between epidemiological Weeks 7 and 17 of 2020, during which parental SARS-CoV-2 strains (WA-1) drove the majority of new cases. Importantly, there were also no baseline differences in the prevalence of anxiety (Fisher’s Exact p = 0.5978 (OR: 1.232 [0.6171 - 2.425]) or depression (p = 0.8339 (OR: 1.192 [0.5394 - 2.607])) across aggregated participant medical histories. Complete demographic features and medical histories are reported in Extended Table 1.

Across all surveyed health dimensions, participants in the Long COVID group demonstrated significant increases in the intensity of reported symptoms and dramatically worsened quality of life (Fig. 1D) (Extended Table 2). To address whether there was a pattern of responses associated with Long COVID, survey responses were aggregated into a single classification metric (Long COVID Propensity Score or LCPS) using a parsimonious logistic regression model (Long COVID vs. Other) which demonstrated both significant diagnostic potential (.939 AUC, bootstrap CI 0.89-0.97) (Fig. 1E-F, Extended Table 3) and enabled subsequent analysis of specific immunological features mediating Long COVID propensity.

Analysis of the prevalence of self-reported symptoms among the Long COVID group revealed frequent reports of fatigue (87%), brain fog (78%), memory difficulty (62%), and confusion (55%) (Fig. 1G), consistent with symptom prevalence reported in numerous prior reports of Long COVID. The prevalence of Postural Orthostatic Tachycardia Syndrome (POTS) was also frequent within the Long COVID group, with 37.6% of individuals having objective clinical diagnoses (**Fig. S1A**). Negative impacts on employment status were also frequently reported by participants within the Long COVID group (51%) (**Fig. S1B**). To further extend analyses of Long COVID symptomology, an agglomerative hierarchical clustering of binary symptom data was performed to identify clusters of participants with similar sets of self-reported symptoms. Three distinct Long COVID clusters were identified (bootstrapped mean cluster-wise Jaccard similarity: Cluster 1: 0.751 [95% CI 0.542-1.00]; Cluster 2: 0.601 [95% CI 0.465-0.944]; Cluster 3: 0.747 [95% CI 0.561-1.00]). Comparison of LCPS across Long COVID clusters demonstrated a clear bifurcation, with Cluster 1 and 2 showing uniformly high propensity scores relative to more moderate LCPS values in Cluster 3 (**Fig. S1C**).

### Immune phenotyping of circulating cell populations from Long COVID participants demonstrated specific elevations in both inflammatory and anti-viral immune responses

Analysis of peripheral blood mononuclear cell populations revealed a significant difference in circulating immune cell populations among MY-LC groups. Levels of non-classical monocytes (CD14^lo^CD16^hi^), traditionally involved in mediating anti-inflammatory activity, as well as vascular homeostasis and viral immune responses ^39–41^, were significantly elevated among Long COVID participants (mean 1.14% LC vs. 0.74% CC vs. 0.86% HC) (Fig. 2A). Abundance of maturing (CD15^+^), MHC Class II (HLA-DR) expressing, non-classical monocytes were similarly elevated (HLA-DR^+^ mean 5.02% LC vs. 1.22% CC vs. 2.60% HC; CD15^+^ mean 30.16% LC vs. 22.45% CC vs. 17.59% HC) (Fig 2A). Significant decreases in circulating populations of cDC1 subsets, typically involved in cross-presentation to CD8^+^ T cells and Th_1_ response polarization^42, 43^, were also noted among Long COVID participants (Fig. 2B). Levels of other circulating granulocyte populations (neutrophils, eosinophils, classical and intermediate monocytes, plasmacytoid dendritic and cDC2 populations) were not significantly different between groups, with significant heterogeneities noted among participants with Long COVID (**Fig. S2A**).

**Figure 2.**
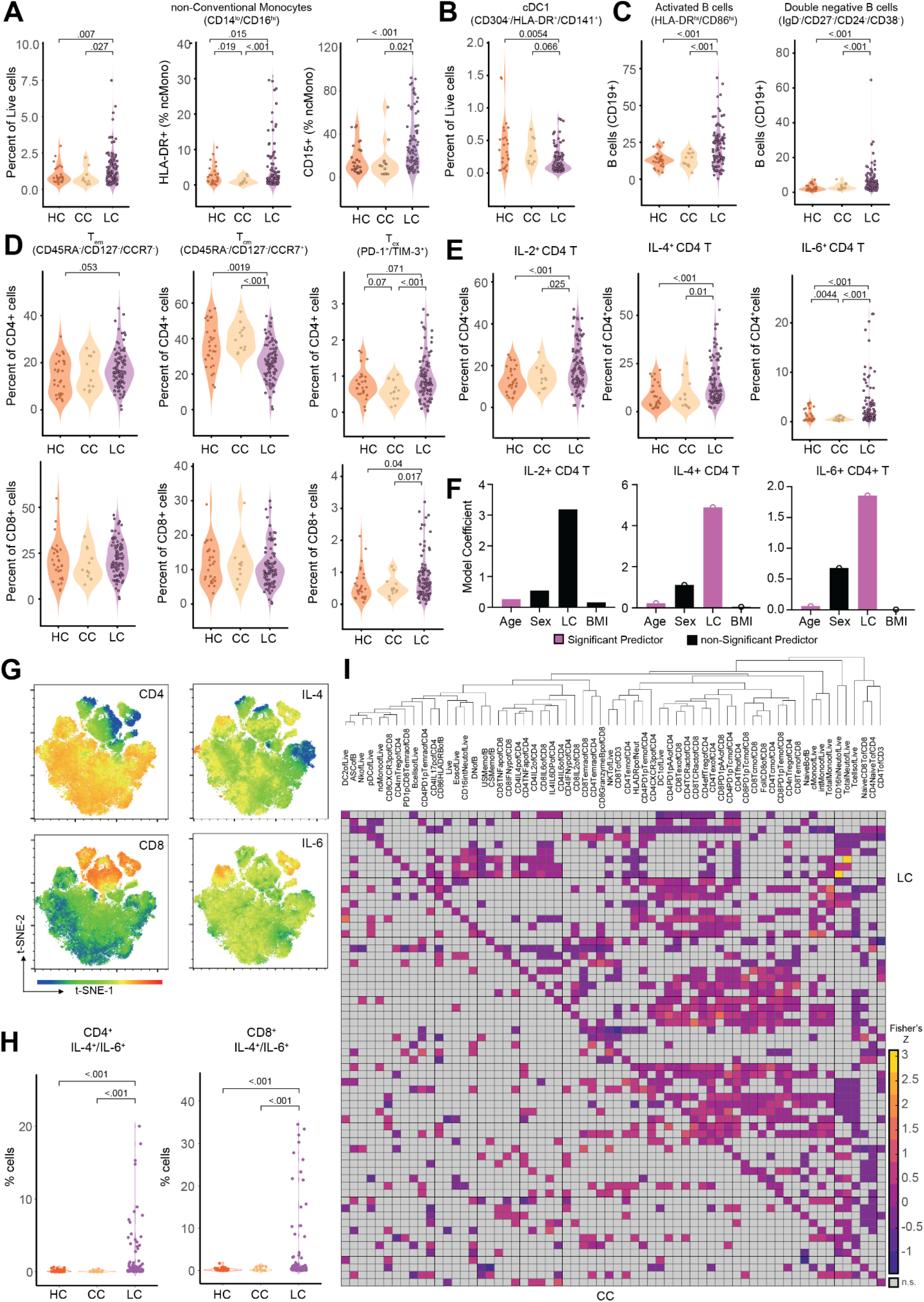
Immunological differences in myeloid and lymphocyte effectors among participants with Long COVID. **(A-B)** Violin plots of myeloid peripheral blood mononuclear populations (PBMCs) plotted as percentages of respective parent populations (gating schemes detailed in Extended Figure 9A). Significance was assessed using one-way ANOVA for differences in group means, with post-hoc analysis using t-tests corrected for multiple corrections using Tukey’s Method. **(C-E)** Violin plots of various lymphocyte peripheral blood mononuclear populations (PBMCs) plotted as percentages of respective parent populations (gating schemes detailed in Extended Figure 9B). Significance was assessed using one-way ANOVA for differences in group means, with post-hoc analysis using t-tests corrected for multiple corrections using Tukey’s Method. **(F)** Model coefficients from generalized linear modeling are reported for various outcomes (top of panels). Significant predictors (p ≤ 0.05) are plotted in purple and non-significant predictors are plotted in black. Detailed model results are reported in Extended Table 5. **(G)** Representative tSNE plots from a participant with Long COVID for various PBMC populations (CD4^+^, CD8^+^, IL-4, and IL-6). Individual dots represent single cells. **(H)** Violin plots of IL-4 and IL-6 double positive CD4^+^ and CD8^+^ T cells plotted as percentages of CD3^+^ cells. Significance was assessed using one-way ANOVA for differences in group means, with post-hoc analysis using t-tests corrected for multiple corrections using Tukey’s Method. **(I)** Hierarchical clustering of Fisher-Z transformed Pearson correlations between various PBMC populations for LC group (upper right triangle) and CC group (lower left triangle). Distance metrics were first calculated using standardized Euclidean metrics with WPGMA linkage among the LC group and resulting dendrogram ordering applied to CC group for direct comparison. For all panels, only significant differences between groups are displayed.

B lymphocyte populations showed increases in both activated populations (CD86^hi^HLA-DR^hi^) (mean 21.09% LC vs. 12.48% vs. 12.87%) and double-negative (IgD^-^/CD27^-^/CD24^-^/CD38^-^) (mean 6.68% LC vs. 2.59% CC vs. 2.53% HC) subsets (Fig. 2C). Circulating levels of naïve B cells and various other B cell subsets were not significantly different across groups (**Fig. S2B**). Flow cytometry analyses of circulating T lymphocyte populations revealed no difference in naïve T cells or effector memory subsets (T_EM_; CD45RA^-^/CD127^-^/CCR7^-^) (**Fig. S2C**) but significant decreases in circulating CD4^+^ central memory (T_CM_; CD45RA^-^/CD127^+^/CCR7^-^) (mean 27.49% LC vs. 33.79% CC vs. 33.51% HC) and significant increases exhausted CD4^+^ and CD8^+^ subsets (T_EX_; PD-1^+^/Tim-3^+^) (CD4_Ex_ mean 0.96% LC vs. 0.77% CC vs. 0.76% HC; CD8_Ex_ average 0.84% LC vs. 0.65% CC vs. 0.62% HC) (Fig. 2D). Analysis of intracellular cytokine production following PMA and ionomycin stimulation showed significant increases in CD4^+^ and CD8^+^ T cell production of IL-2 (CD4 IL-2 mean 19.44% LC vs. 14.20% CC, vs. 13.73% HC; CD8 IL-2 mean 5.86% LC vs. 2.83% CC vs. 2.94% HC) and IL-6 (CD4 IL-6 mean 3.54% LC vs. 1.75% CC, vs. 1.79% HC; CD8 IL-6 mean 5.45% LC vs. 0.82% CC vs. 0.82% HC), and CD4^+^-specific increases in IL-4 production (CD4 IL-4 mean 15.56% LC vs. 8.08% CC, vs. 8.47% HC) (Fig. 2E**, Fig. S2C**). Intracellular levels of TNF-α, IFN-γ, IL-17 (CD4^+^), and GMZB (CD8^+^) were not significantly different across groups (**Fig. S2D-E**). To confirm whether Long COVID status was significantly associated with levels of IL-2, IL-4, and IL-6 after accounting for demographic differences among participants, generalized linear models were constructed incorporating age, sex, Long COVID status (binary), and body mass index (BMI). Both age^44–46^ and Long COVID status were significant predictors of intracellular production of IL-4 and IL-6, but not IL-2 (Fig. 2F, Extended Table 4). Manifold embedding of flow cytometry data from individual participants revealed populations of IL-4^+^/IL-6^+^ double-positive CD4^+^ and CD8^+^ T lymphocytes (Fig. 2G). Subsequent group-wide analysis of IL-4^+^/IL-6^+^ cells in CD4+ and CD8^+^ T cell populations revealed significant increases among participants with Long COVID (CD4 IL-4^+^/IL-6^+^ mean 1.75% LC vs. 0.21% CC vs. 0.23% HC; CD8 IL-4^+^/IL-6^+^ mean 3.74% LC vs. 0.29% CC vs. 0.31% HC), with a significant number of outliers that had upwards of 20-30% double-positive CD4 and CD8 lymphocytes (Fig. 2H). Hierarchical clustering of flow cytometry data within the Long COVID group revealed blocks of co-varying immune effectors (notably T_ex_ and IL-2, IL-4, and IL-6 producing CD4^+^ and CD8^+^ T cells) absent from convalescent controls and consistent with evidence of aberrant, chronic immune engagement among Long COVID participants (Fig. 2I).

### Long COVID participants demonstrated elevated SARS-CoV-2 specific humoral responses

Among participants with two doses of SARS-CoV-2 vaccines and prior infection, there was a significant increase in anti-S1 IgG levels (Fig. 3A). The levels of anti-S and anti-RBD IgG were elevated in LC, but not significantly different from CC (Fig. 3A). Comparison of historical, unvaccinated controls previously infected with SARS-CoV-2 and unvaccinated Long COVID participants demonstrated significant increases in anti-N IgG, while levels of anti-S, anti-S1, and anti-RBD were not significantly different (**Fig. S3A**). To more fully account for differences in demographic factors and differential vaccination status across study groups, statistical modeling accounting for differential number of vaccines at blood draw (Fig. 3B**, Fig. S3B**) and time from most recent vaccination (**Fig. S3C**) revealed that Long COVID status was a significant positive predictor of anti-Spike humoral responses (Extended Table 5). Linear peptide profiling of anti-SARS-CoV-2 IgG responses to Spike revealed elevated levels of binding at known sites conferring increased neutralization potential (amino acid residues (a.a.) 556-572 (1.24x LC vs. CC) & a.a. 572-586 (1.27x))^47, 48^. In addition, enhanced binding was detected at sites enriched in the Long COVID group (a.a. 625-638 (1.65x), a.a. 660-672 (1.38x) (a SARS-CoV-2 infection motif), and a.a. 682-690 (3.05x) (furin cleavage site)) and sites across various S2 residues were enriched in the CC group (a.a. 1149-1161 (1.43x CC vs. LC) & a.a. 1256-1266 (2.65x)) (Fig. 3C). Multiple differentially expressed SARS-CoV-2 Spike binding motifs were mapped onto publicly available structural models of trimeric Spike (PDB 6VXX) and were found to be highly surface exposed in natural conformational states nearby receptor binding domain (RBD) in S1 (RDPQTLE and KFLPQQ), as well as near the S1/S2 cleavage site (RSVAS, YECDIPIGAGICA, and YMSLG), consistent with results of enhanced anti-Spike immune responses among Long COVID participants (Fig. 3D). Specific analysis of individual peptide enrichment for Spike motifs revealed significant increases in the magnitude of humoral responses directed against RDPQTLE (Kruskal-Wallis p = 0.0041) (Fig. 3E).

**Figure 3.**
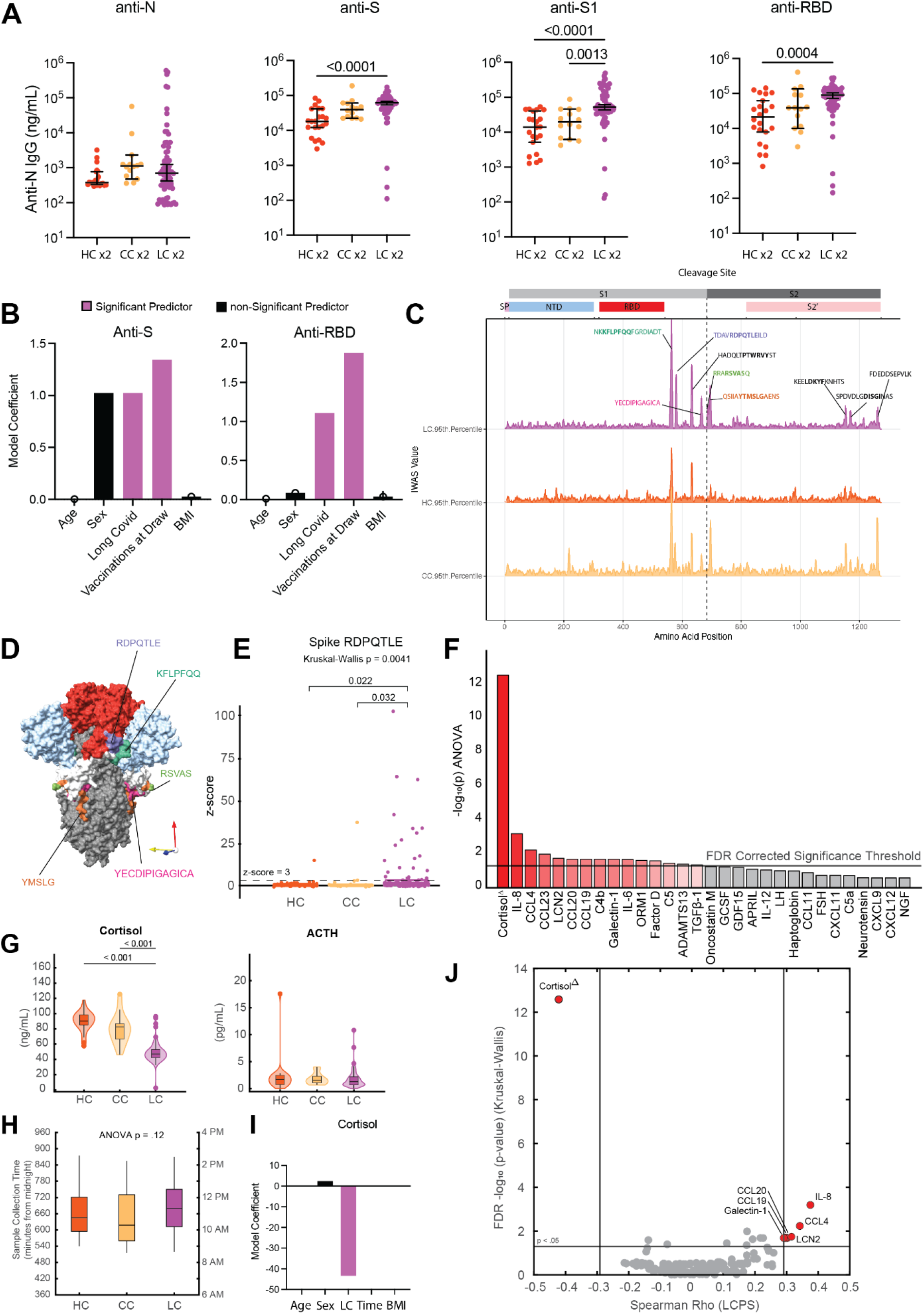
Elevated SARS-CoV-2 specific humoral responses and altered circulating plasma factors among Long COVID participants. **(A)** Dot plots of various anti-SARS-CoV-2 IgG concentrations among vaccination-matched HC, CC, and LC groups (all with prior infection). Vaccination status for each group is indicated by the general form “x2” where digits indicate the total number of SARS-CoV-2 vaccine doses. Central bars indicate group means, and error bars represent 95% confidence interval estimates of group means. Significance for difference in group medians was assessed using Kruskal-Wallis with Dunn’s test for post-hoc comparison. Reported p-values are adjusted for multiple comparison using Tukey’s Method. **(C)** Model coefficients from generalized linear modeling are reported for various outcomes (top of panels). Significant predictors (p ≤ 0.05) are plotted in purple and non-significant predictors are plotted in black. Detailed model results are reported in Extended Table 6. **(D)** IWAS line profiles of IgG antibody binding plotted along SARS-CoV-2 Spike amino acid sequence. Various Spike protein domains are indicated by colored boxes (top). 95^th^ percentile values are displayed for each group: LC (top, purple), HC (middle, orange) and CC (bottom, yellow) with peaks ≥ 2.5 IWAS value annotated by their consensus linear motif sequence (bold) and surrounding residues. **(E)** Three-dimensional mapping of LC-enriched motif sequences onto SARS-CoV-2 trimeric spike protein. (S1 = light grey; N terminal domain (NTD) = light blue; receptor-binding domains (RBD) = red; and S2 = dark grey, with various LC-enriched motifs annotated. **(F)** Box plots of z-score enrichments for IgG binding to SARS-CoV-2 Spike sequence RDPQTLE. A z-score greater than or equal to 3 indicates significant binding relative to SERA control populations. Central lines indicate group medians, with top and bottom lines indicating 75th and 25th percentiles, respectively. Significance was assessed using Kruskal-Wallis with post-hoc comparisons using Dunn’s test corrected from multiple comparison (Tukey’s Method) **(G)** Negative Log_10_ transformed p-values from Kruskal-Wallis tests for various plasma factors are plotted. P-values are corrected for multiple comparison using FDR correction (Benjamini-Hochberg). Only the top 30 factors (ranked by Kruskal-Wallis p-values) are plotted for visualization. Plasma factors with significant differences in group medians are colored in red. “Δ” indicates that participants with potentially confounding medical comorbidities (e.g. cortisol: pre-existing pituitary adenoma, adrenal insufficiency, oral steroid use) were removed prior to analysis for differences in group medians. (**H)** Box-plots of post-hoc comparisons using Dunn’s test are reported, with correction for multiple comparison using Tukey’s Method. Central lines indicate group medians, with top and bottom lines indicating 75th and 25th percentiles, respectively. **(I)** Box-plots of sample collection times are reported. Significant differences in sample collection times were assessed using Kruskal-Wallis with post-hoc comparisons with Dunn’s test. Correction for multiple comparison was performed using Tukey’s Method. Central lines indicate group medians, with top and bottom lines indicating 75th and 25th percentiles, respectively. **(J)** Model coefficients from generalized linear modeling of cortisol levels. Significant predictors (p ≤ 0.05) are plotted in purple and non-significant predictors are plotted in black. Detailed model results are reported in Extended Table 7. **(K)** Bi-plot of plasma factor p-values from Kruskal-Wallis analysis (y-axis) and Spearman correlations with LCPS scores. (x-axis). Horizontal line indicates significance threshold for Kruskal-Wallis analysis. Vertical lines represent the minimum significant correlation values for plasma factors correlating with LCPS scores. Plasma factors exceeding both thresholds are colored in red and labeled.

### Long COVID participants displayed significant perturbations in glucocorticoids and soluble immune mediators

Parallel multiplex analysis of circulating hormones and immune mediators from 99 LC, 15 CC, and 25 HC participant plasma samples revealed significant differences in cortisol (Kruskal-Wallis p = 2.62E-13), IL-8 (Kruskal-Wallis p = 0.000642), CCL4 (Kruskal-Wallis p = 0.00586), CCL23 (Kruskal-Wallis p = 0.0103), LCN2 (Kruskal-Wallis p = 0.0181), CCL20 (Kruskal-Wallis p = 0.0206), CCL19 (Kruskal-Wallis p = 0.0206), C4b (Kruskal-Wallis p = 0.0206), Galectin-1 (Kruskal-Wallis p = 0.0206), IL-6 (Kruskal-Wallis p = 0.0207) and various other soluble factors (Fig. 3F). Post-hoc comparisons of IL-8, CCL4, and other mediators demonstrated significant increases among Long COVID participants (**Fig. S4A-K**). Post-hoc comparison of cortisol concentrations revealed that Long COVID participants had approximately 50% lower circulating levels (median levels 47.01 ng/mL LC vs. 90.32 ng/mL HC vs. 82.67 ng/mL). Evaluation of paired ACTH levels revealed no significant differences across groups (Kruskal-Wallis p = 0.677) (Fig. 3G). Additional analysis of sample collection times revealed no significant differences in collection time (Kruskal-Wallis p = 0.12) (Fig. 3H) and statistical modeling revealed that Long COVID status was a significant predictor of reduced cortisol levels after accounting for individual differences in age, sex, sample collection time, and BMI (Fig. 3I, Extended Table 6). Biplot analysis of p-values resulting from Kruskal-Wallis analysis of circulating concentrations against significant correlations with Long COVID Propensity Scores (LCPS) demonstrated that cortisol was significantly different across MY-LC study groups and negatively correlated with LCPS, whereas levels of galectin-1, CCL19, CCL20, LCN2, CCL4, and IL-8 were significantly different across groups and positively correlated with LCPS (Fig. 3J).

### Long COVID participants did not exhibit increased autoantibodies to the extracellular proteome

Next, antibody reactivity against extracellular human proteins was assessed using Rapid Extracellular Antigen Profiling (REAP), a high throughput method that allows for the measurement of antibody reactivity against >6,000 extracellular and secreted human proteins^21, 49^. Though participants with Long COVID demonstrated a variety of private reactivities against diverse human autoantigens (Fig. 4A), there were no significant differences in the total number of autoantibody reactivities per participant across groups (Fig. 4B), nor was there a significant correlation between the number of reactivities and Long COVID cluster (as assessed by LCPS scores) (Fig. 4C). Additionally, there was also no correlation between number of autoantibody reactivities and double-negative B cell populations or days from acute symptom onset **(Fig. S5A-B)**. Given previous findings of elevated functional autoantibodies in severe acute COVID-19 ^21^, autoantibody reactivities were aggregated into clusters using a manually curated GO Process list relevant to Long COVID. There were no significant differences in the magnitude of reactivity between Long COVID and controls across any of the categories (Fig. 4D).

**Figure 4.**
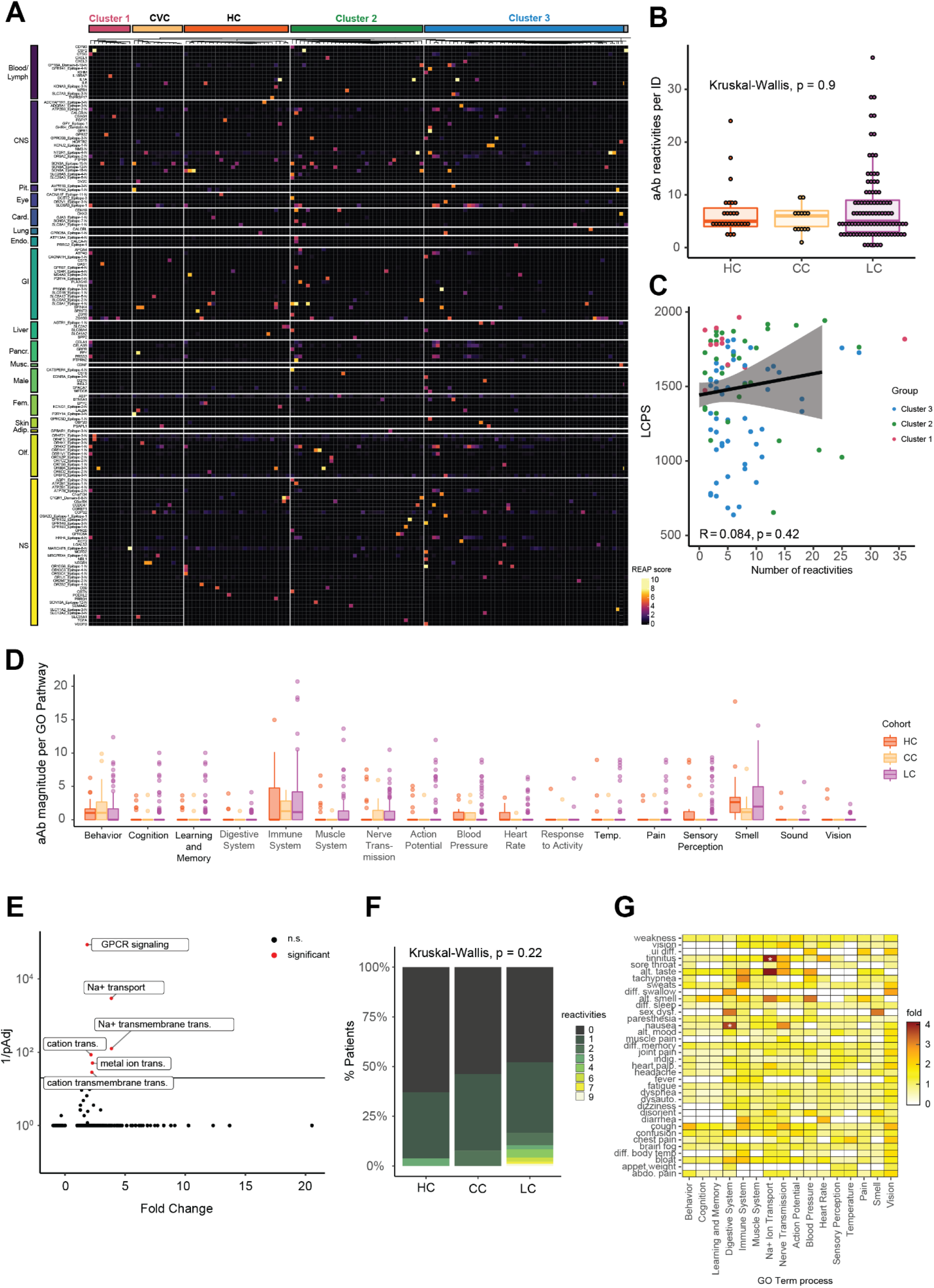
Long COVID participants showed limited but selective autoantibodies against the human exoproteome. **(A)** Heatmap depicting REAP reactivities across the MY-LC Study. Each column is one participant, with participants grouped by study group and LCPS. Column sub-clustering within groups performed by k-means clustering. Each row is one protein. Proteins grouped using Human Protein Atlas mRNA expression data. Reactivities depicted have at least one participant with a REAP score >= 3. **(B)** The number of autoantibody reactivities per individual by group (HC = healthy control; CC = convalescent control; LC = Long Covid). Significance assessed by Kruskal-Wallis. Each dot represents one individual. Boxplot colored box depicts 25th to 75th percentile of the data, with the middle line representing the median. **(C)** Correlation plot depicting the relationship between number of autoantibody reactivities per individual and LCPS. Statistical significance assessed by Spearman correlation. Black line depicts linear regression with 95% CI shaded. Colors depict Long COVID cluster (Cluster 1 = red; Cluster 2 = green; Cluster 3 = blue). Each dot represents one individual. **(D)** Grouped box plot depicting reactivity magnitude per individual in the listed GO Process domain. Reactivity magnitude is calculated as the sum of REAP scores for all reactivities per individual in a given GO Process domain. Statistical significance assessed by Kruskal-Wallis. Boxplot colored box depicts 25th to 75th percentile of the data, with the middle line representing the median, and outliers depicted as individual points. **(E)** Panther GO Biological Process overrepresentation analysis for reactivities unique to Long COVID relative to the background REAP library proteins. Statistical significance determined by Fisher’s exact test with correction for multiple hypotheses by Bonferroni. Red color indicates adjusted p-value <0.05. **(F)** Bar plot depicting the proportion of participants with n reactivities against proteins belonging to the GO Process “sodium ion transport”. Statistical significance assessed by Kruskal-Wallis. **(G)** Heatmap depicting the relationship between binary Long COVID symptoms and reactivity magnitude against a given GO Process domain. Color depicts the fold enrichment in symptom prevalence in individuals at or above the 95th percentile for a given GO Process domain relative to overall prevalence of a given symptom. Statistical significance is marked by asterisks and was assessed by Fisher’s exact test, with correction for multiple comparisons performed using Benjamini-Hochberg method.

Although total levels of autoreactivity were not elevated within the Long COVID group, individual participants could potentially share patterns of autoreactivity that may explain specific disease features. Thus, unbiased analysis of differential antigen targeting in Long COVID and control groups was performed using PANTHER, which revealed unique enrichment in several biological processes related to cation transport, including “sodium ion transport” (GO: 0006814) (Fig. 4E), while both control and Long COVID reactivity lists were enriched for “GPCR signaling pathways” (GO: 0007186) **(Fig. S5C)**. To further explore the unique enrichment in sodium ion transport reactivities, differences were assessed in both the number and magnitude of reactivities against proteins belonging to this GO Process between MY-LC study groups. While the total number (Fig. 4F) and magnitude **(Fig. S5D)** of reactivities were not significantly different across groups, autoantibody reactivities were notably elevated in a subset of participants with Long COVID who displayed reactivities against 6, 7 or even 9 different proteins of this family. Although there was no difference in sodium channel reactivity magnitude between Long COVID clusters (**Fig. S5E**), binarization of Long COVID groups into quantile extremes (95^th^ percentile or greater) for each analyzed GO Process revealed significantly increased incidence of tinnitus (GO Process “sodium ion transport”, p = 0.025) and nausea (GO Process: “digestive system process”, p = 0.034) (Fig. 4G). The individual reactivities driving the sodium ion transporter magnitude were diverse, but most commonly included SLC6A6, sodium-dependent taurine and beta-alanine transporter, ATP1A2, the catalytic component of an Na+/K+ ATPase enriched in the central nervous system and heart, and SLC9A3, a Na+/H+ exchanger. In contrast, differences in Digestive System Process were largely driven by reactivities against Cholecystokinin B receptor (CCKBR), a receptor for Gastrin and Cholecystokinin present in the CNS and GI tract.

### Long COVID participants displayed altered humoral responses to distinct herpesviruses

Given the emerging evidence for a role of latent virus reactivation in Long COVID^37, 50^, anti-viral reactivity patterns were examined in the MY-LC groups. To accomplish this, two complementary approaches were undertaken. Initial analysis of participants’ global anti-viral responses was assessed by Rapid Extracellular Antigen Profiling (REAP), which also measures antibody reactivity to 225 exogeneous viral proteins (Supplementary Table 1). Reactivity against 36 different viral conformational epitopes was detected amongst 99 Long COVID and 40 control participants **(Fig. S6A)**. For comparison of SARS-CoV-2 protein reactivities, analysis was confined to participants with 2 doses of SARS-CoV-2 vaccine. Participants in Long COVID group had significantly elevated mean differences in REAP reactivities against almost all non-Omicron SARS-CoV-2 variant RBD sequences tested relative to healthy and convalescent controls (LC vs. controls mean net-difference: Delta: +2.51 (p = .056); Beta: +2.46 (p = .056); Gamma: +2.33 (p = .014); Alpha: +2.13 (p = .0167; and Epsilon: +1.97 (p = .0298)), in agreement with the ELISA analysis (Fig. 5A, 3A).

**Figure 5.**
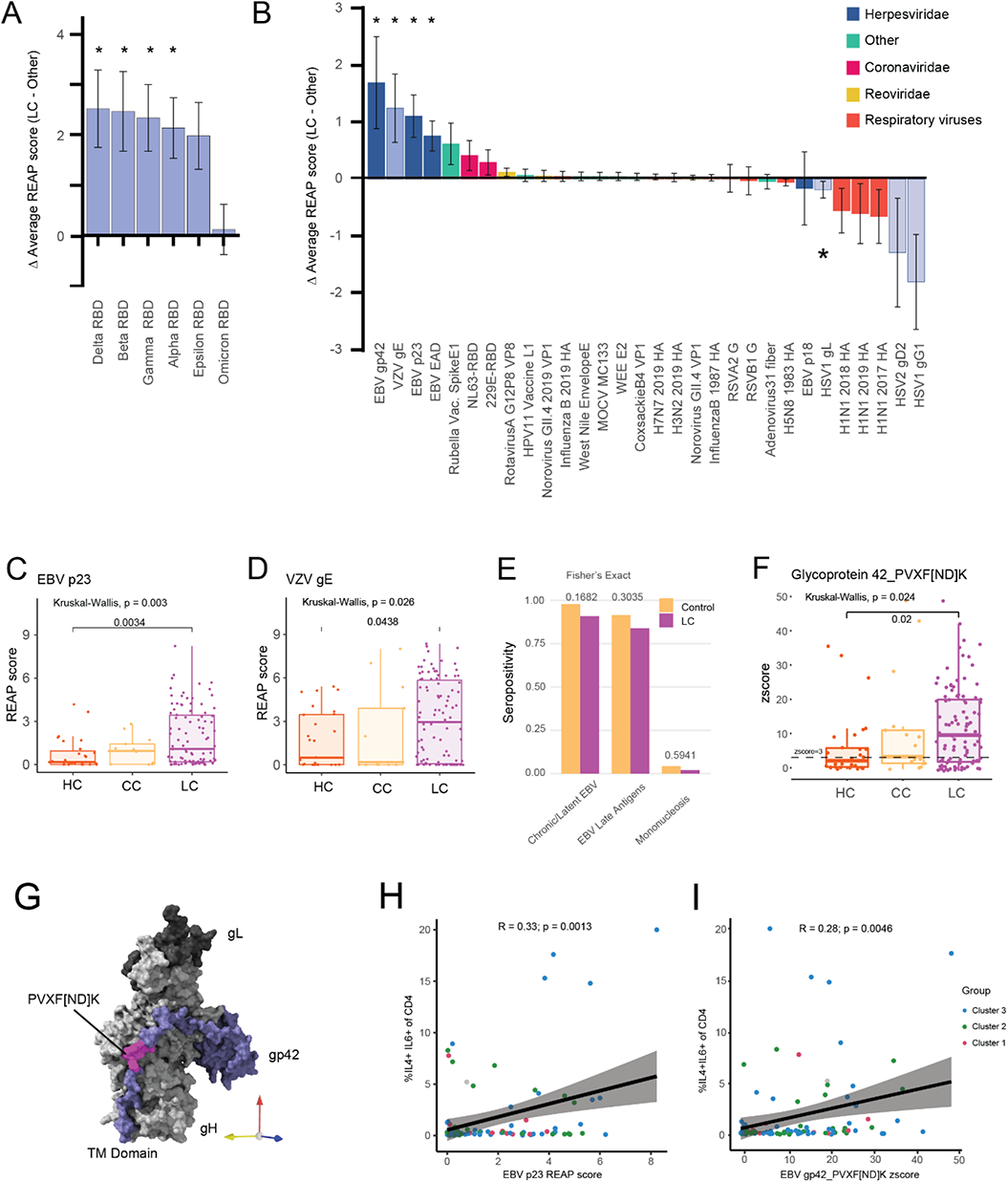
Long COVID participants demonstrate elevated levels of antibody responses to herpesviruses. **(A)** Bar plot depicting the difference in average REAP score for SARS-CoV2 S1 RBD between participants with Long COVID (n = 99) and pooled healthy and convalescent controls (n = 40). Error bars depict the 95% confidence interval. Statistical significance assessed by unpaired Wilcoxon with correction for multiple hypotheses by Benjamini-Hochberg method. **(B)** Bar plot depicting the difference in average REAP score for a given viral antigen between Long COVID participants (n = 99) and pooled healthy and convalescent controls (n = 40). Error bars depict the 95% confidence interval. Statistical significance assessed by unpaired Wilcoxon with correction for multiple hypotheses by Benjamini-Hochberg method. **(C, D)** REAP scores for EBV p23 (5C) and VZV gE (5D) by group (HC = healthy control; CC = convalescent control; LC = Long COVID). Statistical significance determined by Kruskal Wallis. Post-hoc tests performed using Dunn’s test with Holm’s method to adjust for multiple comparisons. Each dot represents one individual. Boxplot colored box depicts 25th to 75th percentile of the data, with the middle line representing the median. **(E)** Proportion of each group seropositive for three EBV disease panels (Chronic/Latent EBV, EBV Late Antigens, and Mononucleosis) as determined by SERA (LC = Long COVID). Statistical significance determined by Fisher’s Exact test. **(F)** SERA-derived z-scores for EBV gp42 motif PVXF[ND]K plotted by group (HC = healthy control; CC = convalescent control; LC = Long Covid). Statistical significance determined by Kruskal Wallis. Post-hoc tests performed using Dunn’s test with Holm’s method to adjust for multiple comparisons. Each dot represents one individual. Boxplot colored box depicts 25th to 75th percentile of the data, with the middle line representing the median. Dashed line represents z-score threshold for epitope positivity defined by SERA. (**5G**) Three-dimensional mapping of LC-enriched linear peptide sequence PVXF[ND]K (magenta) onto EBV gp42 (purple) in complex with gH (light grey) and gL (dark grey) (PDB: 5T1D). **(H, I)** Correlation plot depicting the relationship between EBV p23 REAP score **(H)** or EBV gp42 PVXF[ND]K z score **(I)** and %IL4+/IL6+ of CD4+. Statistical significance assessed by Pearson correlation. Black line depicts linear regression with 95% CI shaded. Colors depict Long COVID cluster (Cluster 1 = red; Cluster 2 = green; Cluster 3 = blue). Each dot represents one individual.

Differences in mean viral reactivities against non-SARS-CoV2 antigens were also striking (Fig. 5B), with participants in the Long COVID group displaying elevated mean REAP scores against several herpesvirus antigens including the Epstein-Barr Virus (EBV) minor viral capsid antigen gp23 (Fig. 5C), the EBV fusion receptor component gp42 **(S6B),** the EBV acute phase antigen EAD **(Fig. S6C),** and the VZV glycoprotein E (Fig. 5D). To orthogonally validate the REAP findings, ELISAs for EBV p23 **(Fig. S6D)** and EBV EAD **(Fig. S6E)** were performed, finding a significant correlation between REAP scores and corresponding ELISA reactivities.

To further interrogate herpesvirus-specific antibody responses among each of the groups, the Serum Epitope Repertoire Analysis (SERA) platform was used. SERA is a commercially available random bacterial display library which encompasses linear epitope panels representing 45 different pathogens and disease markers, previously validated leveraging a database of thousands of controls^51^. Importantly, this analysis revealed no significant difference in the estimated seroprevalence of EBV across study groups (91% EBV seropositive Long COVID participants vs. 98% seropositive controls, Fisher’s Exact Test p = .1682) (Fig. 5E) or any other viral pathogens (**Fig. S6F**). By linear peptide screening, reactivity against EBV-associated epitopes was similar within the MY-LC study groups, though Long COVID participants had higher seropositivity for two linear motifs mapping to two envelope glycoproteins, gp42 and gp350, both essential for lytic infection of B-cells by EBV **(Fig. S6G).** Moreover, the Long COVID group had a higher degree of reactivity against the gp42 linear peptide, PVXF[ND]K (Fig. 5F). This motif was mapped onto publicly available structural models of gp42 in complex with EBV gH/gL (PDB 5T1D) and the identified residues were found to be highly surface exposed in its natural conformational state on the surface of EBV virions (pink residues, Fig. 5G).

Aggregating the initial results of REAP and SERA, Long COVID individuals had higher titers of anti-EBV antibodies, even though overall seroprevalence is not different from healthy or convalescent controls.

Additional analysis revealed no statistically significant differences between LCPS and humoral reactivity directed against EBV p23 or gp42 PVXF[ND]K antigens **(Fig. S6H, S6I).** In contrast, the reactivity to both of these herpesvirus antigens correlated with populations of activated T-cells, including IL4/IL6 producing CD4+ T cells in Long COVID participants (EBV p23: r = 0.34, gp42 PVXF[ND]K: p = 1.1E-3; r = 0.28, p = 4.6E-3) (Fig. 5H, 5I). Furthermore, additional significant correlations were observed between EBV p23 reactivity and terminally differentiated effector memory (T_EMRA_) CD4 T cells (r = 0.45, p = 7.4E-6) **(Fig. S6J),** a subset of cells previously implicated in protection from CMV^72, 73^ and Dengue virus^74^. In contrast, there was no correlation between anti-SARS-CoV-2 antibody levels and IL-4/IL-6 double-positive CD4 T cells (**Fig. S7**).

### Unsupervised and supervised machine learning approaches identified unique biological markers of Long COVID

UMAP embedding of study participants with all collected immunological features demonstrated a clear visual separation between people with Long COVID and those without (Fig. 6A). Consistent with this, k-nearest neighbors classification on the normalized features demonstrated efficient discriminative performance, with an AUC=0.96 (95% CI: 0.92-0.99, Fig. 6B). Principal components regression of collated immunological data identified flow cytometry (pseudo-R^2^ = 73.0%) and plasma proteomics (pseudo-R^2^ = 64.6%) as the most informative individual data blocks contributing to efficient separation of groups, whereas autoantibodies against human exoproteome (aAb) contributed the least. A final parsimonious LASSO model similarly achieved good fit (pseudo-R^2^ = 91.3%) (Fig. 6C). Of the features selected for the final model, several features significantly distinguished Long COVID (elevated double negative B cells, serum galectin-1 concentration, various EBV epitopes) while others were negatively associated (reduced serum cortisol, PD-1^+^ CD4^+^ T_CM_, and HSV1 and HSV2 motifs) (Fig. 6D).

**Figure 6.**
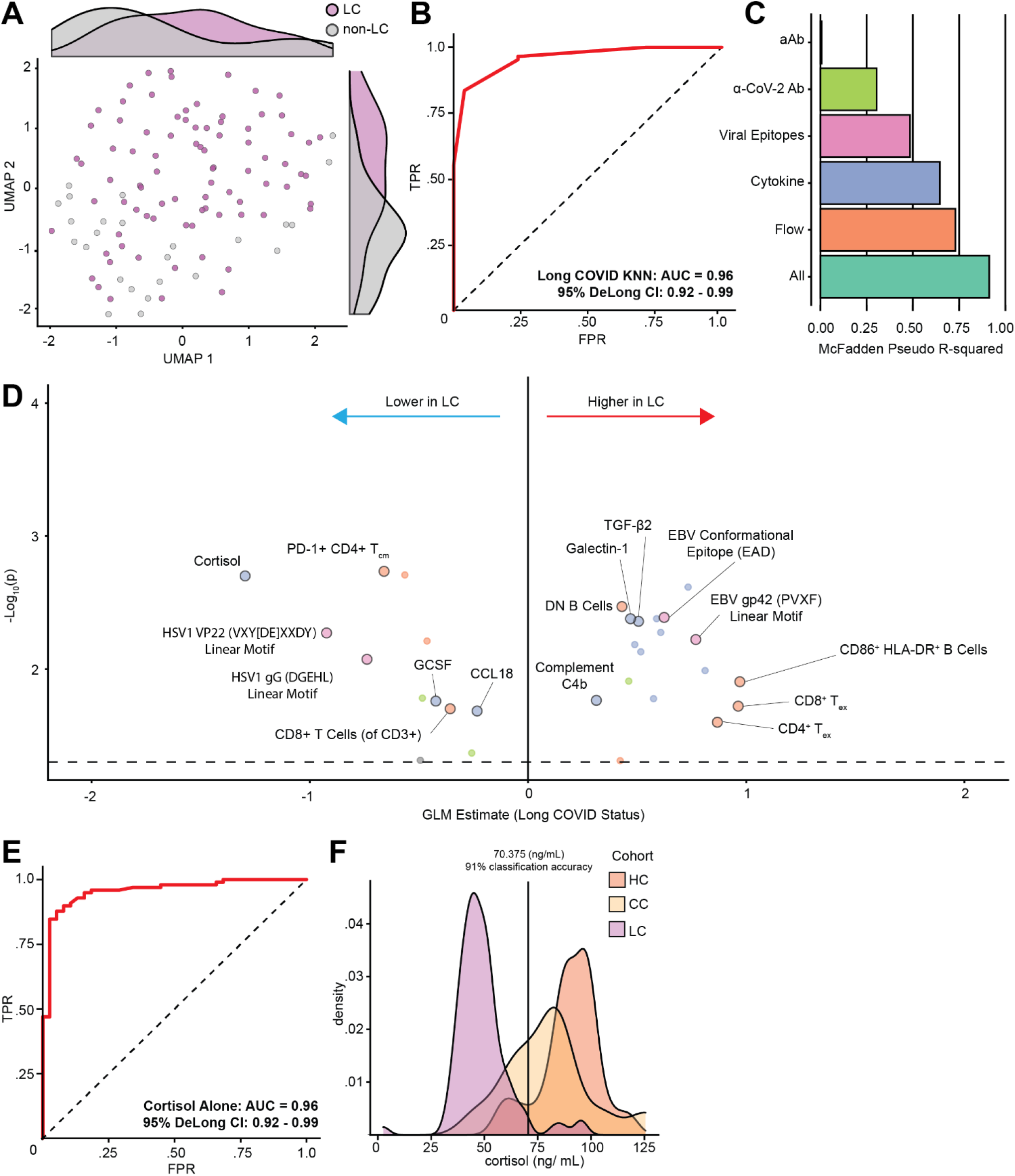
Biochemical factors differentiate participants with Long COVID from controls. **(A)** UMAP projection of participant data comprised of cytokine, flow cytometry, and various antibody responses (SARS-CoV-2-specific, non-SARS-CoV-2 viral antibodies, and autoantibodies). Marginal histograms display data density along each UMAP dimension. **(B)** ROC curve analysis from unsupervised K-nearest neighbors (KNN) classification. AUC and 95% CI intervals (DeLong’s Method) are reported. **(C)** McFadden’s pseudo R-squared are reported as bar plot for each data segment. An integrated, parsimonious McFadden’s pseudo R-squared is reported for the final classification model (‘All’). **(D)** LASSO regression identifies minimal set of immunologic features differentiating participants with Long COVID from others. Unlabeled dots are significant predictors not included in the final LASSO regression model. Dots are colored according to individual data segments: orange = Flow cytometry, blue = Plasma cytokines, pink = viral epitopes, green = SARS-CoV-2 specific antibodies, yellow = autoantibodies to human exoproteome (aAb). **(E)** ROC curve analysis using cortisol levels as an individual classifier of Long COVID status. AUC and 95% CI intervals (DeLong’s Method) are reported. **(F)** Kernel-density smoothed histograms of cortisol for HC, CC and LC groups. Vertical line depicts threshold values of cortisol with maximal discriminatory accuracy. *Abbreviations: LC = Long COVID; TPR = true positive rate; FPR = false positive rate; KNN = k-nearest neighbors; AUC = area under the curve; CI = confidence interval; aAb = autoantibodies to human exoproteome; α-SARS-CoV-2 = anti-SARS-CoV-2 antibodies; Flow = flow cytometry*.

Serum cortisol was the most significant individual predictor of Long COVID status in the model, and cortisol alone as a predictor achieved an AUC of 0.96 (95% CI: 0.92-0.99) in the data set (Fig. 6E). Notably, serum cortisol within the MY-LC study was highest in healthy (uninfected) controls, lower in convalescent controls, and lowest in participants with Long COVID (Fig. 6F). When tuning for accuracy, a threshold of 70.38 ng/mL obtained a maximum classification accuracy of 91.9% (Fig. 6F). Comparison of classification accuracies between LCPS models (Fig. 1 E-F) and machine learning (Fig. 6B) revealed substantial agreement (Cohen’s Kappa .865, 95% CI [0.83 -.90]), suggesting that both PROs and immunological features efficiently classify Long COVID (Extended Table 7).

To assess the association between immune profiling and Long COVID propensity, the original UMAP of immunological features was used to visually assess separation of Long COVID participants according to LCPS (**Fig. S8A**). There was modest capacity for prediction of LCPS scores based on k-nearest neighbors (AUC = 0.68, 95% CI: 0.58-0.78) as compared to classification of Long COVID status (**Fig. S8B**). PCA regression model construction demonstrated moderate fit of the integrated model (pseudo-R^2^ = 71.9%) and once again identified flow cytometry (pseudo-R^2^ = 55.7%) and plasma proteomics (pseudo-R^2^ = 71.3%) as the most informative data segments (**Fig. S8C**). Serum cortisol was again the principal significant factor negatively associated with LCPS in the model (**Fig. S8D**).

## Discussion

Persistent sequelae are a prominent and debilitating consequence of infection with SARS-CoV-2^1, 3, 29^. Our exploratory analyses identified key significant immunological differences relative to demographically matched control populations at >400 days post infection. A number of significant changes in circulating leukocytes, including increases in non-classical monocytes, activated B cells, double-negative B cells, exhausted T cells, and IL-4/IL-6 secreting CD4 T cells, and decreases in conventional DC1 and central memory CD4 T cells were identified. In addition, antibodies to SARS-CoV-2 antigens and herpesvirus lytic antigens were elevated in participants with Long COVID. In contrast, no significant differences were found for autoantibodies to human exoproteome. Most strikingly among participants with Long COVID, levels of plasma cortisol were roughly half of those found in healthy or convalescent controls. Based on machine learning, cortisol levels alone were the most significant predictor for Long COVID classification, as well as for estimation of Long COVID Propensity Score. Multiple hypotheses have been proposed for Long COVID pathogenesis, including persistent virus/virus remnants, autoimmunity, dysbiosis, latent viral reactivation and unrepaired tissue damage ^18, 32–38^. Our data suggest the involvement of persistent antigen, reactivation of latent herpesviruses, and chronic inflammation, and are less consistent with the autoantibodies to extracellular antigens.

Immune phenotyping of PBMC populations revealed notable elevations in circulating non-classical monocytes among the Long COVID group. Non-classical monocytes are frequently associated with anti-inflammatory responses programs; however, they are also engaged in maintenance of vascular homeostasis, Th_1_ anti-viral response polarization, regulation of immune complex deposition, and various chronic inflammatory and autoimmune conditions ^39–41, 52^. Significant decreases in levels of circulating cDC1 populations were also observed among participants with Long COVID, which are classically associated with antigen presentation and cytotoxic T cell priming during viral infection ^57, 58^. In parallel with perturbations in circulating myeloid populations, significant reductions in the CD4^+^ T_CM_ cells and elevations in exhausted CD4^+^ and CD8^+^ T cells were observed. Analysis of intracellular cytokine production following stimulation displayed significant increases in the production of inflammatory IL-2 and IL-6 among CD4^+^ and CD8^+^ T cells, and a specific elevation of IL-4 CD4^+^ T cells. Unexpectedly, subsets of Long COVID participants also had polyfunctional IL-4 / IL-6 co-expressing CD4^+^ T cells which correlated with antibody reactivity against EBV lytic antigens, but not SARS-CoV-2 antigens. In aggregate, these findings are consistent with chronic immune engagement against reservoirs of viral antigen among participants with Long COVID.

With respect to humoral immunity, SARS-CoV-2 specific IgG against Spike and S1 were elevated in Long COVID participants as compared to vaccination-matched controls. Linear peptide profiling of antibody binding across SARS-CoV-2 Spike demonstrated both exaggerated magnitude and unique binding targets among participants with Long COVID, most notably at residues 682-690 comprising the furin cleavage site. These findings are consistent with results from analysis of PBMC populations, such as T_EX_ increases suggesting chronic immune responses directed against viral antigens within the Long COVID group. Intriguingly, circulating Spike protein has also been observed in Long COVID participants, but not in convalescent controls ^53^. Furthermore, these findings are supported by prior reports of persistent viral antigen in intestinal biopsies of convalescent COVID-19 individuals, and suggest persistent antigen might drive the continuous elevation in antibody responses among people with Long COVID^33–36^.

In parallel to the analysis of humoral responses and circulating immune effector populations, systematic profiling of soluble immune mediators found numerous significant differences among MY-LC study groups. Participants with Long COVID demonstrated striking decreases in systemic cortisol levels – decreases which remained significant after accounting for differences in individual demographic factors and sample collection times. This hypocortisolism was not associated with a significant perturbation in ACTH levels, suggesting an inappropriately blunted compensatory response by the hypothalamic-pituitary axis. The significance of the magnitude and prevalence of hypocortisolism in individuals with Long COVID is highlighted in that low levels of cortisol were also the dominant feature driving the accurate separation of Long COVID participants in machine learning models. Prior reports have associated low cortisol levels during the early phase of COVID-19 in patients that develop respiratory Long COVID symptoms^37^. Thus, our current finding of *persistently* decreased cortisol production in participants with Long COVID more than a year following acute infection warrants expanded investigation. Importantly, cortisol plays a critical role in mediating homeostatic stress responses and hypocortisolism shares substantial clinical overlap with Long COVID symptoms^54^. Low levels of cortisol have also been reported for myalgic encephalomyelitis/chronic fatigue syndrome (ME/CFS)^55^ and treatment with hydrocortisone is reported to elicit modest improvement in symptoms. However, adrenal suppression has ultimately precluded its widespread clinical use for this indication^55^ and additional clinical trials may be needed to optimize glucocorticoid replacement therapies for Long COVID and ME/CFS.

The multi-dimensional immune profiling of Long COVID participants also revealed elevated humoral immune responses to non-SARS-CoV-2 viral antigens, particularly EBV. EBV viremia was previously reported during acute COVID-19^37, 56^ in hospitalized patients and predicted the development of persistent symptoms in the post-acute COVID-19 period ^37^. The observation of elevated IgG against EBV lytic antigens in this study suggests recent reactivation of latent herpesviruses (EBV and VZV) may be a common feature of Long COVID. Additionally, concordant analysis of EBV IgG responses by REAP and SERA found significant positive correlations between reactivity against EBV p23 and T_EMRA_ and IL- 4^+^/IL-6^+^ CD4 T cell populations, as well as correlations between reactivity against EBV gp42 and IL-4^+^/IL-6^+^ double positive CD4 T cell populations among participants with Long COVID. These results suggest that herpesvirus reactivation is not incidental following SARS-CoV-2 infection, and instead that non-SARS-CoV-2 viral pathogens may alternatively mediate, aggravate, or exploit the persistent changes observed in circulating immune effector populations. Whether EBV reactivation may also predispose people with Long COVID to the development or exacerbation of autoimmune pathologies, as has been recently reported for people with multiple sclerosis^57, 58^, will require extensive longitudinal monitoring and surveillance of people with Long COVID.

Extensive autoantibody profiling indicated that there were no stereotypical or shared extracellular autoantibodies that could differentiate participants with Long COVID from controls. Furthermore, there was no correlation between the degree of autoantibody reactivity and Long COVID propensity, nor was there any disproportionate targeting of functional pathways to distinguish Long COVID. In context of prior hypotheses suggesting that autoantibodies may contribute to the pathogenesis of Long COVID^18, 38^, our results suggest they play a more limited role in disease pathology. However, the present results suggest autoantibodies may associate with particular features of Long COVID symptoms in subsets of affected individuals, such as tinnitus and nausea. Given the generally sparse, private nature of the detected autoreactivities, a prospective study with additional statistical power will be required to validate and detect more subtle patterns in autoantibody reactivity amongst participants with Long COVID that could play a disease-modifying role. Additionally, whether autoantibodies may be associated with other adverse clinical outcomes following COVID-19 merits future study.

Finally, machine learning models identified multiple significant predictors of Long COVID status relative to convalescent and healthy control populations. While cortisol was the most robust individua predictor of Long COVID status, maintaining its excellent specificity for Long COVID diagnoses outside of the MY-LC study is unlikely given its known pleiotropic role in a variety of diverse disease processes. Instead, it is proposed that a minimal set of soluble biomarkers identified in this study (decreased cortisol, increased IL-8 and galectin-1) may serve as more specific diagnostic biomarkers for Long COVID. Additionally, classification accuracy using solely immunological data obtained from Long COVID participants strongly agreed with Long COVID classifications using LCPS scoring (Cohen’s Kappa – 0.865), demonstrating that both PROs and immunological analyses are highly concordant in diagnosing Long COVID.

Importantly, this study has several limitations. Primary among these considerations is the relatively small number of participants that were extensively immunophenotyped. While broad in its coverage of diverse biological features, this study lacked the thousands of independent observations that traditional machine learning methods rely upon to robustly train and optimize classification models. Instead, this study leveraged machine learning to parse hundreds of individuals, each with tens of thousands of data points, to identify a suite of candidate immunological features important in the accurate classification of Long COVID. Naturally this approach limits broad applicability without external validation, and the reported observations should serve primarily to guide and inform future studies investigating mediators of Long COVID pathogenesis. Beyond sample size, this study also prioritized analysis of peripheral (circulating) immune factors from study participants. As Long COVID often presents with organ-system specific dysfunctions, a greater emphasis on analysis of local - as well as systemic - immune features would serve as a critical extension of the findings presented in the current study. Further, our analysis of autoantibodies was restricted only to the exoproteome. Whether antibodies to intracellular autoantigens and non-protein autoantigens play a role in Long COVID pathogenesis was not tested. Lastly, analysis of cortisol levels among participants also displayed excellent specificity for a Long COVID diagnosis within the MY-LC study; however, the reported accuracies do not account for other disease processes that phenotypically resemble Long COVID or other disease processes where hypocortisolism is prominent.

In summary, significant biological differences have been identified between participants with Long COVID and demographically and medically matched convalescent and healthy control groups, validating the extensive reports of persistent symptoms by various Long COVID advocacy groups. Unbiased machine learning models further identified both putative biomarkers of Long COVID, as well as potential mediators of Long COVID disease pathogenesis. Our study provides a basis for future investigations into the immunological underpinnings driving the genesis of Long COVID.

## Data Availability

All data produced in the present study are available upon reasonable request to the authors.

## Methods

### Ethics Statement

This study was approved by the Mount Sinai Program for the Protection of Human Subjects (IRB #20-01758) and Yale Institutional Review Board (IRB #2000029451 for MY-LC; IRB #2000028924 for enrollment of pre-vaccinated Healthy Controls). Informed consent was obtained from all enrolled participants.

### MY-LC Study Design, Enrollment Strategy, and Inclusion / Exclusion Criteria

MY-LC was a cross-sectional, multi-site study comprised of four different groups with differing SARS-COV-2 exposure histories and varied Long COVID status. The HC, CC and LC groups had samples collected within the Mount Sinai Healthcare System. The HCW group had samples collected within the Yale New Haven Healthcare System. Participants enrolled in the Long COVID group previously underwent complete medical evaluations by physicians to rule out alternative medical etiologies for their persistent symptoms before study enrollment.

Participants with persistent symptoms following acute COVID-19 were recruited from Long COVID clinics within the Mount Sinai Healthcare System and the Center for Post COVID care at Mount Sinai Hospital. Participants enrolled in healthy and convalescent study arms were recruited via IRB-approved advertisements delivered through email lists, study flyers located in hospital public spaces, and on social media platforms. Informed consent was provided by all participants at the time of enrollment. All participants provided peripheral blood samples and completed symptom surveys the day of sample collection (described below). Self-reported participant medical histories were also collected at study visits and corroborated via review of electronic medical records. Prevalence of specific medical conditions was estimated through synthesis of self-reports and retrospective review of EMR data, where either participant self-reports or the presence of the condition in the EMR constituted a positive result for the condition.

Inclusion criteria for the Long COVID group were age ≥ 18 years; previous confirmed or probable COVID-19 infection (according to World Health Organization guidelines^59^); and persistent symptoms > 6 weeks following initial COVID-19 infection. Inclusion criteria for enrollment of healthy controls were age ≥ 18 years, no prior COVID-19 infection, and completion of a brief, semi-structured verbal screening with research staff confirming no active symptomatology. Inclusion criteria for convalescent controls were age ≥ 18 years; previous confirmed or probable prior COVID-19 infection; and completion of a brief, semi-structured verbal screening with research staff confirming no active symptomatology.

Pre-specified exclusion criteria for this study were inability to provide informed consent; and any condition preventing a blood test from being performed. Additionally, all participants had electronic health records reviewed by study clinicians following enrollment and were subsequently excluded prior to analyses for the following reasons: (1) current pregnancy, (2) immunosuppression equivalent to or exceeding prednisone 5 mg daily, (3) active malignancy or chemotherapy, and (4) any monogenic disorders. For specific immunological analyses, pre-existing medical conditions were additionally excluded prior to analyses due to high potential for confounding (e.g., participants with hypothyroidism were excluded prior to analysis of circulating T3/T4 levels; participants with pituitary adenomas were excluded prior to analysis of cortisol levels). Specific exclusions are marked by “Δ” in figures and detailed in relevant legends.

### Participant Surveys

A comprehensive suite of surveys was administered to MY-LC study participants, combining validated patient-reported outcomes (PROs) with custom, purpose-developed tools by the MY-LC study team. Baseline demographic data collected from surveys included gender, age, body mass index (BMI), race, and medical comorbidities. Additionally, participants in the Long COVID and convalescent group were asked to provide COVID-19 clinical data including date of symptom onset and acute disease severity (non-hospitalized vs. hospitalized), any SARS-CoV-2 polymerase chain reaction (PCR) diagnostic testing results, and any SARS-CoV-2 antibody testing results, Finally, all participants were asked to report SARS-CoV-2 vaccination status including date of vaccinations and vaccine brand.

At the time of blood collection, all participants completed patient-reported outcomes (PROs) for fatigue (Fatigue Severity Scale^60^, fatigue visual analogue scale [F-VAS]), post-exertional malaise (DePaul Symptom Questionnaire Post-Exertional Malaise Short Form [DSQ-PEM Short Form])^61^, breathlessness (Medical Research Council [MRC] Breathlessness Scale^62^), cognitive function (Neuro-QOL v2.0 Cognitive Function Short Form^63^), health-related quality of life (HRQoL) (EuroQol EQ-5D-5L^64^), anxiety (GAD-7^65^), depression (PHQ-2^66^), pain (P-VAS), sleep (Single-Item Sleep Quality Scale^67^), and pre- and post-COVID-19 employment status (author-developed). Lastly, participants in the MY-LC study were asked to self-report any current persistent symptoms from a study provided list.

All survey data were collected and securely stored using REDCap^68, 69^ (Research Electronic Data Capture) electronic data capture tools hosted within the Mount Sinai Health System.

### Long COVID Propensity Scoring (LCPS)

Evaluation of immunophenotyping results with an objectively derived propensity score was achieved through construction of a multivariable logistic regression model generated with Long COVID vs. others (Healthy Controls + Convalescent controls) as the outcome. Predictor variables included FSS^60^, F-VAS, DSQ-PEM Short Form, MRC Breathlessness Scale^62^, Neuro-QOL v2.0 Cognitive Function Short Form^63^, EQ-5D-5L^64^, GAD-7^65^, PHQ-2^66^, P-VAS, Single-Item Sleep Quality Scale^67^. Model selection using Akaike’s Information Criteria (AIC) was used to select the final, parsimonious model. Log odds values from the final model were normalized by dividing them by their respective standard error (SE) and rounding to the nearest integer. These integer values were considered the score items for these specific variables and a cumulative propensity score for each subject was calculated by summation (*Equation 1*, below). As the score did not significantly differ between healthy controls and convalescent controls, the two control groups were combined as a single group (“others”) for final analysis. A ROC curve analysis was performed to identify the optimal cutoff for the LCPS score using the maximum value of Youden’s index J for Long COVID vs others. A 10-fold cross-validation was used for internal validation and to obtain 95% confidence interval (CI) for the area under the curve (AUC). Data were analyzed using Stata version 16 (StataCorp, College Station, Texas).

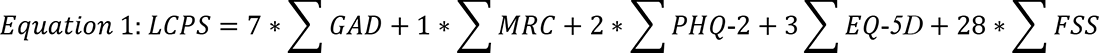

### Blood Sample Processing

Whole blood was collected in sodium-heparin-coated vacutainers (BD 367874, BD Biosciences) from participants at Mount Sinai Hospital in New York City, New York. Following blood draw, all participant samples were assigned unique MY-LC study identifiers and de-identified by research staff. Samples were couriered directly to Yale University in New Haven, CT the same day as the sample collection. Blood samples were processed the same day as collection. Plasma samples were collected after centrifugation of whole blood at 600×g for 10 minutes at room temperature (RT) without brake. Plasma was then transferred to 15-mL polypropylene conical tubes, aliquoted, and stored at 80°C. The peripheral blood mononuclear cell (PBMC) layer was isolated according to the manufacturer’s instructions using SepMate tubes (StemCell). Cells were washed twice with phosphate-buffered saline (PBS) before counting. Pelleted cells were briefly treated with ACK lysis buffer (ThermoFisher) for 2 minutes and then counted. Viability was estimated using standard Trypan blue staining and a Countess II automated cell counter (ThermoFisher). PBMCs were stored at −80°C for cryopreservation or plated directly for flow cytometry studies.

### Flow cytometry

Freshly isolated PBMCs were plated at 1–2 × 10^6^ cells per well in a 96-well U-bottom plate. Cells were resuspended in Live/Dead Fixable Aqua (ThermoFisher) for 20 min at 4 °C. Cells were washed with PBS and followed by Human TruStain FcX (BioLegend) incubation for 10 min at RT. Cocktails of staining antibodies were added directly to this mixture for 30 minutes at RT. Prior to analysis, cells were washed and resuspended in 100 μl 4% PFA for 30 min at 4 °C. For intracellular cytokine staining following stimulation, the surface marker-stained cells were resuspended in 200 μl cRPMI (RPMI-1640 supplemented with 10% FBS, 2 mM L-glutamine, 100 U/ml penicillin, and 100 mg/ml streptomycin, 1 mM sodium pyruvate) and stored at 4 °C overnight. Subsequently, these cells were washed and stimulated with 1× Cell Stimulation Cocktail (eBioscience) in 200 μl cRPMI for 1 h at 37 °C. Fifty μl of 5× Stimulation Cocktail in cRPMI (plus protein transport 442 inhibitor, eBioscience) was added for an additional 4 hours of incubation at 37 °C. Following stimulation, cells were washed and resuspended in 100 μl 4% paraformaldehyde for 30 min at 4 °C. To quantify intracellular cytokines, cells were permeabilized with 1× permeabilization buffer from the FOXP3/Transcription Factor Staining Buffer Set (eBioscience) for 10 min at 4 °C. All subsequent staining cocktails were made in this buffer. Permeabilized cells were then washed and resuspended in a cocktail containing Human TruStain FcX (BioLegend) for 10 min at 4 °C. Finally, intracellular staining cocktails were added directly to each sample for 1 h at 4 °C. Following this incubation, cells were washed and prepared for analysis on an Attune NXT (ThermoFisher). Data were analyzed using FlowJo software version 10.8 software (BD). Antibody information can be seen in Supplementary Table 2. T-distributed stochastic neighbor embedding (t-SNE) visualization of flow cytometry data was performed using FlowJo.

### SARS-CoV-2 antibody testing by ELISA

ELISA assays were performed as previously described ^22^. Briefly, Triton X-100 and RNase A were added to plasma samples at final concentrations of 0.5% and 0.5 mg/ml, respectively, and incubated at room temperature for 30 minutes prior to use to reduce the risk of any potential infectious virus in plasma. MaxiSorp plates (96 wells; 442404, Thermo Scientific) were coated with 50 μl per well of recombinant SARS-CoV-2 Total ectodomain S trimer (SPN-C52H9-100 μg, ACROBiosystems), RBD (SPD-C52H3-100 μg, ACROBiosystems) and the nucleocapsid protein (NUN-C5227-100 μg, ACROBiosystems) at a concentration of 2 μg/ml in PBS and were incubated overnight at 4 °C. The coating was removed and plates were incubated for 1 hour at room temperature with 200 μl of blocking solution (PBS with 0.1% Tween-20 and 3% milk powder). Plasma was diluted serially at 1:100, 1:200, 1:400 and 1:800 in dilution solution (PBS with 0.1% Tween-20 and 1% milk powder), and 100 μl of diluted serum was added for 2 hours at room temperature. Human anti-spike (SARS-CoV-2 human anti-spike [AM006415; 91351, Active Motif]) and human anti-nucleocapsid (SARS-CoV-2 anti-nucleocapsid [1A6; MA5-35941, Active Motif]) were serially diluted to generate a standard curve. Plates were washed three times with PBS-Tween (PBS with 0.1% Tween-20) and 50 μl of HRP anti-human IgG antibody (1:5,000; A00166, GenScript) added to each well in dilution solution. After 1 hour of incubation at room temperature, plates were washed six times with PBS-Tween. Plates were developed with 100 μl of TMB Substrate Reagent Set (555214, BD Biosciences) and the reaction was stopped after 5 min by the addition of 2N sulfuric acid. Plates were then read at an excitation/emission wavelength of 450 nm and 570 nm.

### Multiplex proteomic analysis

Participant plasma was isolated and stored at −80°C as described above. Plasma was shipped to Eve Technologies (Calgary, Alberta, Canada) on dry ice and analytes were measured using the following panels: Human Cytokine/Chemokine 71-plex Discovery Assay (HD71), Steroid/Thyroid 6plex Discovery Assay (STTHD), TGF-Beta 3-plex Discovery Assay (TGFβ1-3), Human Myokine Assay (HMYOMAG-10), Human Neuropeptide Assay (HNPMAG-05), Human Pituitary Assay (HPTP1), Human Cytokine P3 Assay (HCYP3-07), Human Cytokine Panel 4 Assay (HCYP4-19), Human Immunoglobulin Assay (HGAM301-06), Human Adipokine Panel 2 Assay (HADK2-03), Human Cardiovascular Disease Panel Assay (HDCVD9), Human CVD2 Assay (HCVD2-8), Human Complement Panel Assay (HDCMP1), Human Adipokine Assay (HDADK5).

### Linear Peptide Profiling (Serimmune)

#### SERA serum screening

A detailed description of the SERA assay has been published^51^. For this study, plasma was incubated with a fully random 12-mer bacterial display peptide library (1 × 10^10^ diversity, 10-fold oversampled) at a 1:25 dilution in a 96-well, deep well plate format. Antibody-bound bacterial clones were selected with 50 µL Protein A/G Sera-Mag SpeedBeads (GE Life Sciences, #17152104010350) (IgG). The selected bacterial pools were resuspended in growth media and incubated at 37 °C shaking overnight at 300 RPM to propagate the bacteria. Plasmid purification, PCR amplification of peptide-encoding DNA and barcoding with well-specific indices was performed as described. Samples were normalized to a final concentration of 4 nM for each pool and run on the Illumina NextSeq500. Every 96-well plate of samples processed for this study contained healthy control run standards to assess and evaluate assay reproducibility and possible batch effects.

#### PIWAS analysis

The published PIWAS method^70^ was used to identify antigen and epitope signals against the Uniprot reference SARS-CoV-2 proteome (UP000464024). For each sample, approximately 1–3 million 12-mers are obtained from the SERA assay and these are decomposed into constituent 5- and 6-mers. Enrichment scores for each k-mer are calculated by dividing the number of unique 12-mers containing the k-mer divided by the number of expected k-mer reads for the sample, based on amino acid proportions in the sample. The PIWAS analysis was run on the IgG SERA data with a single sample per participant (versus 1500 discovery pre-pandemic controls) and 1010 validation controls were used as the normalization group. 95th quantile bands were calculated based on each population separately.

#### IMUNE-based motif discovery

Peptide motifs representing epitopes or mimotopes of SARS CoV-2-specific antibodies were discovered using the IMUNE algorithm^71^. A total of 164 antibody repertoires from 98 hospitalized subjects from the Yale IMPACT study^22^ were used for motif discovery. The majority of subjects were confirmed SARS CoV-2 positive by NAT. IMUNE compared ∼30 disease repertoires with ∼30 pre-pandemic controls and identified peptide patterns that were statistically enriched (*p*-value ≤ 0.01) in ≥25% of disease and absent from 100% of controls. Multiple assessments were run with different subsets of cases and controls. Peptide patterns identified by IMUNE were clustered using a point accepted mutation 30 (PAM30) matrix and combined into motifs. The output of IMUNE included hundreds of candidate IgG and IgM motifs. A motif was classified as positive in a given sample if the enrichment was ≥3 times the standard deviation above the mean of the training control set. The candidate motifs were further refined based on at least 98% specificity. The final set of motifs was validated for sensitivity and specificity on an additional 1500 pre-pandemic controls and 406 unique confirmed COVID-19 cases from four separate cohorts.

#### Motif grouping by similarity

For SARS-CoV-2, motifs were grouped if they shared at least 3 of 5 amino acid identities, resulting in 76 motifs being assigned into 24 groups. The motif within an epitope group with the greatest sensitivity and mean enrichment was included in the SARS-CoV-2 Infection IgG panel results. In some cases, two motifs were selected from the same group since their combination improved sensitivity. The remaining motifs that did not fall into a group were further down-selected based on a specificity of >99.5%, leaving 24 additional motifs.

### Rapid Extracellular Antigen Profiling (REAP)

#### REAP Library Expansion

The initial yeast library (Exo201) was generated as previously described^21, 49^. In Exo201, only extracellular domains >49 amino acids in length were included in the library. This library was expanded by supplementing expression diversity with the addition of all extracellular domains of multi-pass membrane proteins greater than 15 amino acids and also added 225 diverse viral antigens. Larger antigens that were omitted in Exo201 were additionally backfilled into the library. DNA for new antigens was synthesized as either a Gene Fragment (for antigens over 300 nucleotides) or as an Oligo pool by TWIST Bioscience, containing a 5’ sequence (CTGTTATTGCTAGCGTTTTAGCA) and 3’ sequence (GCGGCCGCTTCTGGTGGC) for PCR amplification. The oligo pool was PCR amplified and transformed into yeast with barcode fragments, followed by barcode-antigen pairing identification as previously described^21, 49^. This new yeast library was then pooled with the initial library (Exo201) in the ratio of 1:1 to generate the new version of the library (Exo205) which contained 6,452 unique antigens.

#### REAP analysis

Participant IgG isolation and REAP selections were performed as previously described^21, 49^. Briefly, IgG was purified from participant plasma using protein G magnetic beads followed by adsorption to yeast transformed with the pDD003 empty vector to remove yeast-reactive IgG. The Exo205 yeast library was induced in SGO-Ura medium and 10^8^ induced yeast cells were washed with PBE and added to wells of a sterile 96-well plate. 10 μg of purified participant IgG was added to the yeast library in duplicate in 100 μL PBE and incubated for 1 hour at 4C. Yeast cells were washed with PBE and incubated with 1:100 biotin anti-human IgG Fc antibody (clone QA19A42, Biolegend) for 30 minutes. Yeast cells were washed with PBE and incubated with a 1:20 dilution of Streptavidin MicroBeads (Miltenyi Biotec) for 30 minutes. Yeast were resuspended in PBE and IgG-bound yeast were isolated by positive magnetic selection using the MultiMACS M96 Separator (Miltenyi Biotec) according to manufacturer instructions and as previously described^21, 49^. Selected yeast were resuspended in 1 mL SDO -Ura and incubated at 30 ⁰C for 24 hours and then harvested for NGS analysis. NGS library preparation was performed as previously described^21, 49^. Briefly, DNA was extracted from yeast libraries using Zymoprep-96 Yeast Plasmid Miniprep kits or Zymoprep Yeast Plasmid Miniprep II kits (Zymo Research) according to standard manufacturer protocols. A first round of PCR was used to amplify a DNA sequence containing the protein display barcode on the yeast plasmid. A second round of PCR was performed on 1 µL step 1 PCR product using Nextera i5 and i7 dual-index library primers (Illumina). PCR products were pooled, run on a 1% agarose gel, and DNA corresponding to the band at 257 base pairs was cut. DNA (NGS library) was extracted using a QIAquick Gel Extraction Kit (Qiagen) according to standard manufacturer protocols. NGS library was sequenced using an Illumina NextSeq550 and an NextSeq high output sequencing kit with 75 base pair single-end sequencing according to standard manufacturer protocols. Approximately 500,000 reads (on average) per sample was collected and the pre-selection library was sampled at ten times greater read depth than other samples. Samples with less than 50,000 reads were classified as a sequencing failure and removed from further analysis.

#### Data analysis

REAP scores were calculated as previously described^21, 49^. Briefly, barcode counts were extracted from raw NGS data using custom codes and counts from technical replicates were summed. Next, aggregate and clonal enrichment was calculated using edgeR^72^ and custom computer scripts. Aggregate enrichment is the log_2_ fold change of all barcodes associated with a particular protein summed in the post-library relative to the pre-library, with zeroes in the place of negative fold changes. Log_2_ fold change values for clonal enrichment were calculated in an identical manner, but barcode counts across all unique barcodes associated with a given protein were not summed. Clonal enrichment for a given reactivity was defined as the fraction of clones out of total clones that were enriched (log_2_ fold change ≥ 2). Aggregate (E_a_) and clonal enrichment (E_c_) for a given protein, a scaling factor (β_u_) based on the number of unique yeast clones (yeast that have a unique DNA barcode) displaying a given protein, and a scaling factor (β_f_) based on the overall frequency of yeast in the library displaying a given protein were used as inputs to calculate the REAP score, which is defined as follows:

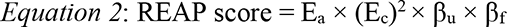

β_u_ and β_f_ are logarithmic scaling factors that progressively penalize the REAP score of proteins with low numbers of unique barcodes or low frequencies in the library, and are described in detail in previous publications^21, 49^.

Antigens with an average REAP score greater than 0.5 across all samples were defined as non-specific and excluded from further analysis. Autoantibody reactivities were defined as antigens with REAP score greater than or equal to 1.

### Statistical Analysis

Study sample was not pre-determined through formal power analysis. Specific statistical methodology can be found in relevant figure legends and manuscript text. Generally, comparison of immunophenotypic features including systemic cytokine levels and antibody concentrations between study groups was performed using non-parametric Kruskal-Wallis tests. In cases where Kruskal-Wallis testing indicated significant differences, post-hoc testing using Dunn’s test was performed with Tukey’s correction for multiple comparison. All statistical tests were performed using R, PRISM, and MATLAB software.

Hierarchical clustering of flow cytometry populations was initially performed among the Long COVID groups using Fisher-Z transformed Pearson R correlations (Z = atanh(R)) with standardized Euclidean distances and WPGMA linkages to generate PBMC population clusters. Dendrogram ordering derived from the Long COVID group was then applied to data from CC group and a composite matrix of transformed correlations generated for visualization.

### Machine Learning

#### Data Preprocessing

All collected data for immune profiling were collated. Features containing redundant information were manually removed from the dataset (e.g., nested flow cytometry populations include only the extant population).

All features were linearly scaled to unit variance and zero-centered using the R programming language base libraries^73, 74^. Median absolute deviation was calculated for each feature across all samples, with missing values removed. Features with a median absolute deviation equal to zero, or features where data was not available in at least half the samples were not included in downstream analysis.

#### Unsupervised Analysis

Principal component analysis, as well as uniform manifold approximation and projection (UMAP), were performed on the set of normalized features^75^. To assess how well participants were grouped by all features, a k-nearest neighbor classifier with k=10 was applied to two scenarios: (1) separating participants with Long COVID from those without (either convalescent participants or healthy controls), and (2) separating participants with extreme LCPS values from other individuals. In the second scenario, participants were classified as having extreme LCPS values if the participant had an LCPS equal to or above the 80^th^ percentile for the dataset. Area under the receiver operating characteristic curve (AUC) and 95% confidence intervals were calculated using DeLong’s method; p-values were calculated using the Mann-Whitney U statistic^76, 77^.

#### Supervised Analysis

Principal components regression was applied to each of a predefined set of data segments: autoantibodies, SARS-CoV-2 antibodies, non-SARS-CoV-2 viral antibodies, plasma proteomics, and flow cytometry readouts. The precise definitions of these data segments are provided as metadata. The first *n* principal components based on explained variance (see below for selection method) were selected from the normalized feature set and used to fit a generalized linear model with two configurations, either binomial with a logit link (equivalent to logistic regression) or gaussian with an identity link (equivalent to ordinary least squares regression). A binomial model with a logit link function was used to determine factors driving classification of participants with Long COVID as compared to convalescent participants without long term symptoms and uninfected controls. A gaussian model with an identity link function was used to determine factors associated with Long COVID Propensity scores (LCPS).

To determine the optimal value for *n* (number of principal components), values were scanned and seven-fold cross validation was performed on the data set. The average mean squared error was calculated for each cross-validation iteration at a particular value of *n*. For the binomial regression run using a logit link function, McFadden’s pseudo-R^2^ was calculated and averaged across each of the cross-validation folds. For the gaussian regression run using an identity link function, a standard R^2^ was calculated.

Plots of explained variance and mean squared error across all scanned values for *n* were generated and visually inspected to choose an optimal value for *n* that maximized explained variance while minimizing overfitting as identified by increasing average mean squared error.

In relating a model fitted on the first *n* principal components to each of the original features, each principal component may be considered as a weighted linear combination of the original features. The principal component loading vectors were used to project the fitted beta values from the generalized linear model using the linearity of expectation, E(X + Y) = E(X) + E(Y), such that the estimated parameter for each variable was the weighted sum of the parameter estimates for the principal components to which it contributed. The variance of fit for each of the original features was similarly projected from the fitted principal components as the variance of a sum of random variables Var(X + Y) = Var(X) + Var(Y) + 2Cov(X, Y). P-values were calculated for each variable in the original feature space using z-scores.

Following per-segment model construction and evaluation, features with a Bonferroni-corrected p-value of less than 0.05 were selected for inclusion in a final principal components regression. These selected features were considered as a separate integrated data segment and processed in the same way as each individual data segment. A least absolute shrinkage and selection operator (LASSO) regression was employed to select a subset of the features with p-values less than 0.05 as a minimal model, and McFadden’s pseudo-R^2^ was calculated.

An implementation has been made publicly accessible as an R library on GitHub at (https://github.com/rahuldhodapkar/puddlr).

#### Symptom Bi-clustering

Participants with Long COVID were clustered based on binary self-reporting of Long COVID symptoms. Hamming distance was used with complete linkage clustering as an agglomeration method. Visualization of the bi-clustering was performed using the ComplexHeatmap package in R^78^. Cluster stability was assessed by bootstrapped resampling with 100 iterations using the fpc package in R^79^.

### Data availability

All the background information on HCWs, clinical information for patients, and raw data used in this study are included in Supplementary Table 1. Additionally, all of the raw fcs files for the flow cytometry analysis are available at ImmPort (TBD).

### Code availability

All computer codes are available as indicated in *Methods* (e.g. github) or available upon request.

## Acknowledgements

The MY-LC study team would like to extend profound thanks to all study participants for donating time, biospecimens, and critical insights on the various health impacts of Long COVID. We also thank M. Linehan for technical and logistical assistance. Various graphical schematics were created with BioRender.com. Lastly, this work was supported by grants from National Institute of Allergy and Infectious Diseases (R01AI157488 to A.I.), FDA Office of Women’s Health Research Centers of Excellence in Regulatory Science and Innovation (CERSI) (to A.I.), Fast Grant from Emergent Ventures at the Mercatus Center (to A.I.), RTW Foundation (to D.P.), the Howard Hughes Medical Institute Collaborative COVID-19 Initiative (to R.M. and A.I.), and the Howard Hughes Medical Institute (to A.I. and R.M.).

## Author contributions

Experimental conceptualization, methodology, and data visualization were performed by J.K., J.W., J.J. P.L., R.D., J.G., A.T., A.A.M., K.K., K.G., V.M., M.P., S.O.; formal analysis conducted by J.K., J.J., P.L. R.D., A.T., and A.A.M.; resources provided by D.V.D., A.M.R., D.P., and A.I.; clinical review of electronic health records was performed by J.K., J.W., J.G. and L.T.; sample collection, processing, and biospecimen validation were performed by J.K.; J.W., J.J., P.L., J.G., A.T., L.T., V.M., M.P., T.M., B.B., T.K., C.L., J.S., D.M., E.B., J.T.M., K.A., T.J.Z., L.X.; writing – original draft by J.K. and A.I.; writing – review & editing by J.K., J.W., J.J. P.L., R.D., J.G., A.T., A.A.M., K.K., H.K., D.V.D., A.M.R., D.P., and A.I.; data curation by J.K., J.J., R.D.; supervised by D.V.D., A.M.R., D.P. and A.I.; funding acquisition by R.M., D.V.D., A.M.R., D.P. and A.I.

## Additional Information

**Supplementary information** is included within this manuscript as Supplementary Tables 1 and 2.

**Correspondence and requests for materials** should be addressed to: A.I., D.P., A.M.R., and D.V.D.

## Extended Figures

**Extended Figure S1.**
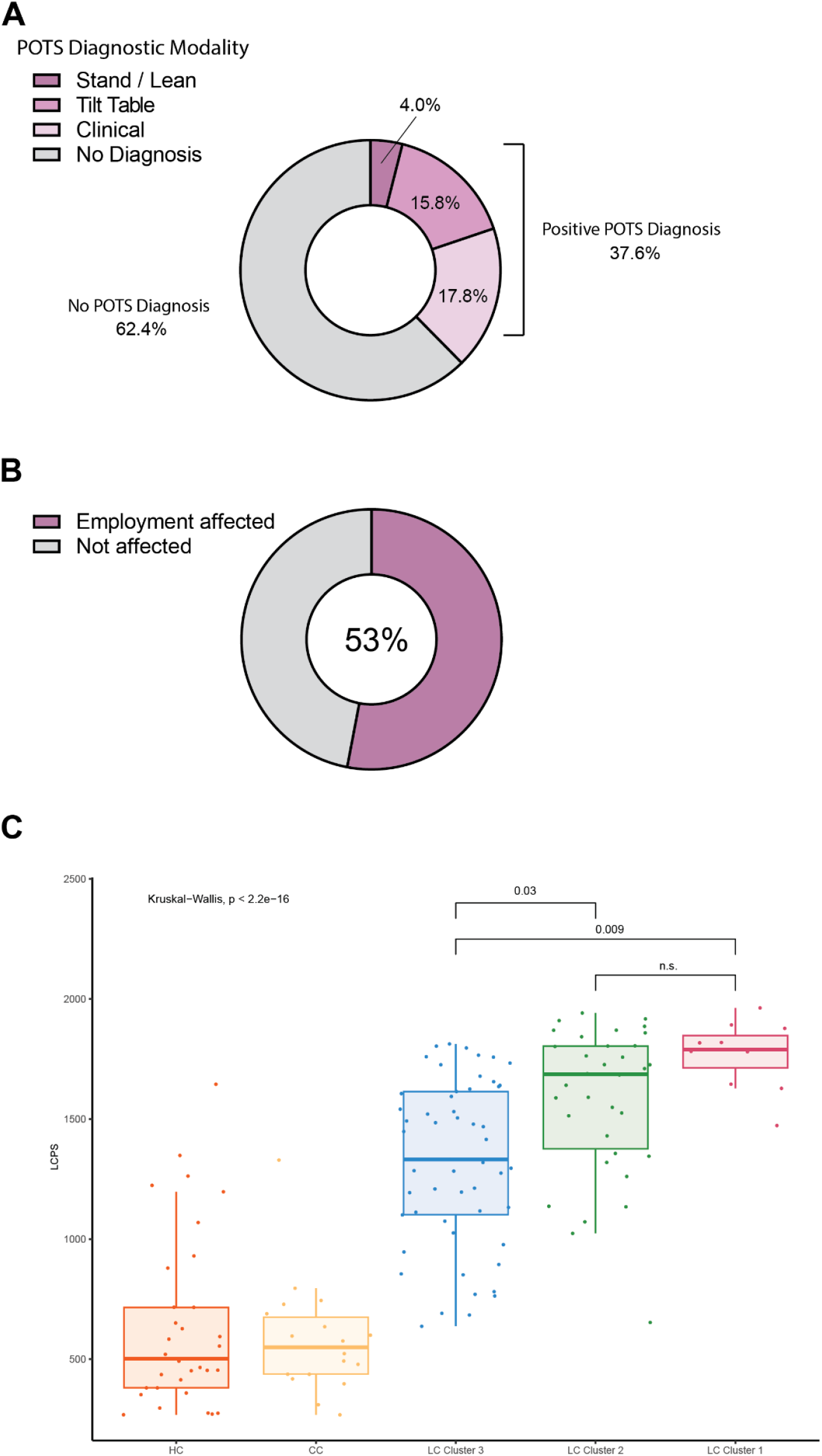
Additional demographic and clinical analysis of the MY-LC Long COVID group. **(A)** Ring plots of prevalence of Postural Orthostatic Tachycardia Syndrome (POTS) among Long COVID group. No diagnosis is represented by grey regions, positive diagnosis is represented by purple regions. Positive POTS diagnoses are further stratified by diagnostic modality: clinical = POTS diagnosed through clinical evaluation (light purple); Tilt-table = POTS diagnosed by Tilt-table (middle purple); Stand / Lean = POTS diagnosed by Stand / LEAN test (dark purple). **(B)** Ring plots of prevalence of self-reported negative impacts on employment status among Long COVID group. Reports of “Not affected” are represented by grey region, reported positive for negative impact are indicated by purple region. **(C)** Box-plots of LCPS across MY-LC study groups. Central colored lines represent group medians, top and bottom lines represent 75^th^ and 25^th^ percentiles respectively. Significance for difference in median LCPS was assessed using Kruskal-Wallis with Dunn’s test for post-hoc comparison. Reported p-vales are corrected using Tukey’s method.

**Extended Figure S2.**
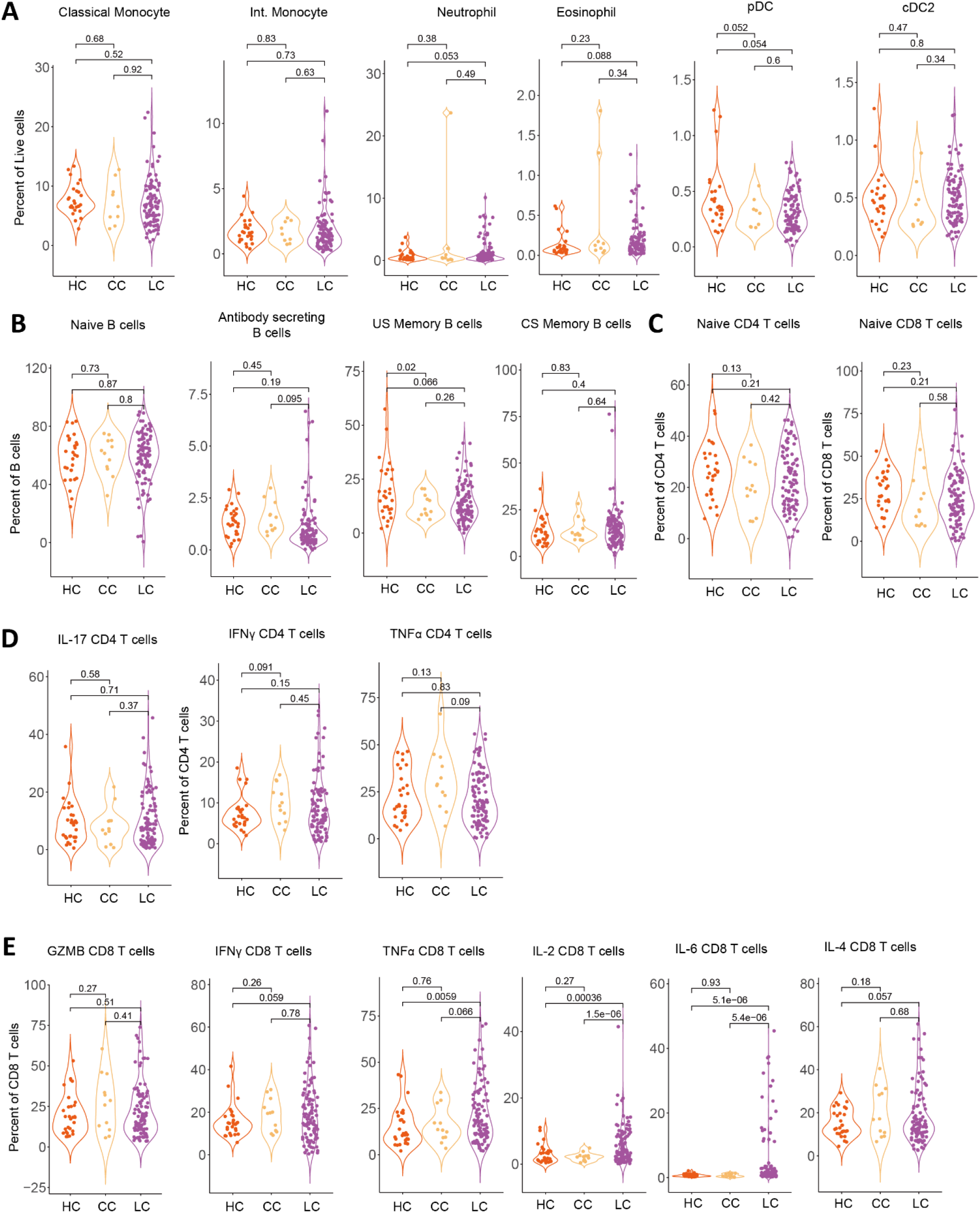
Circulating myeloid, B cell, naïve T cell, and cytokine producing immune cell populations among MY-LC participants. **(A-B)** Violin plots of various myeloid and B cell PBMC populations across healthy (HC), convalescent (CC), and Long COVID (LC) groups. Significance for difference in group averages was assessed using ANOVA with two-sample t-tests for post-hoc comparison and shown where significant. Reported p-vales are corrected using Tukey’s method. **(C)** Violin plots of naïve CD4 and CD8 T cell populations across healthy (HC), convalescent (CC), and Long COVID (LC) groups. Significance for difference in group averages was assessed using ANOVA with two-sample t-tests for post-hoc comparison and p-values shown between groups. Reported p-vales are corrected using Tukey’s method. **(D-E)** Violin plots of intracellular cytokine production following PMA/Ionomycin stimulation across healthy (HC), convalescent (CC), and Long COVID (LC) groups. Significance for difference in group averages was assessed using ANOVA with two-sample t-tests for post-hoc comparison and p-values shown between groups. Reported p-vales are corrected using Tukey’s method.

**Extended Figure S3.**
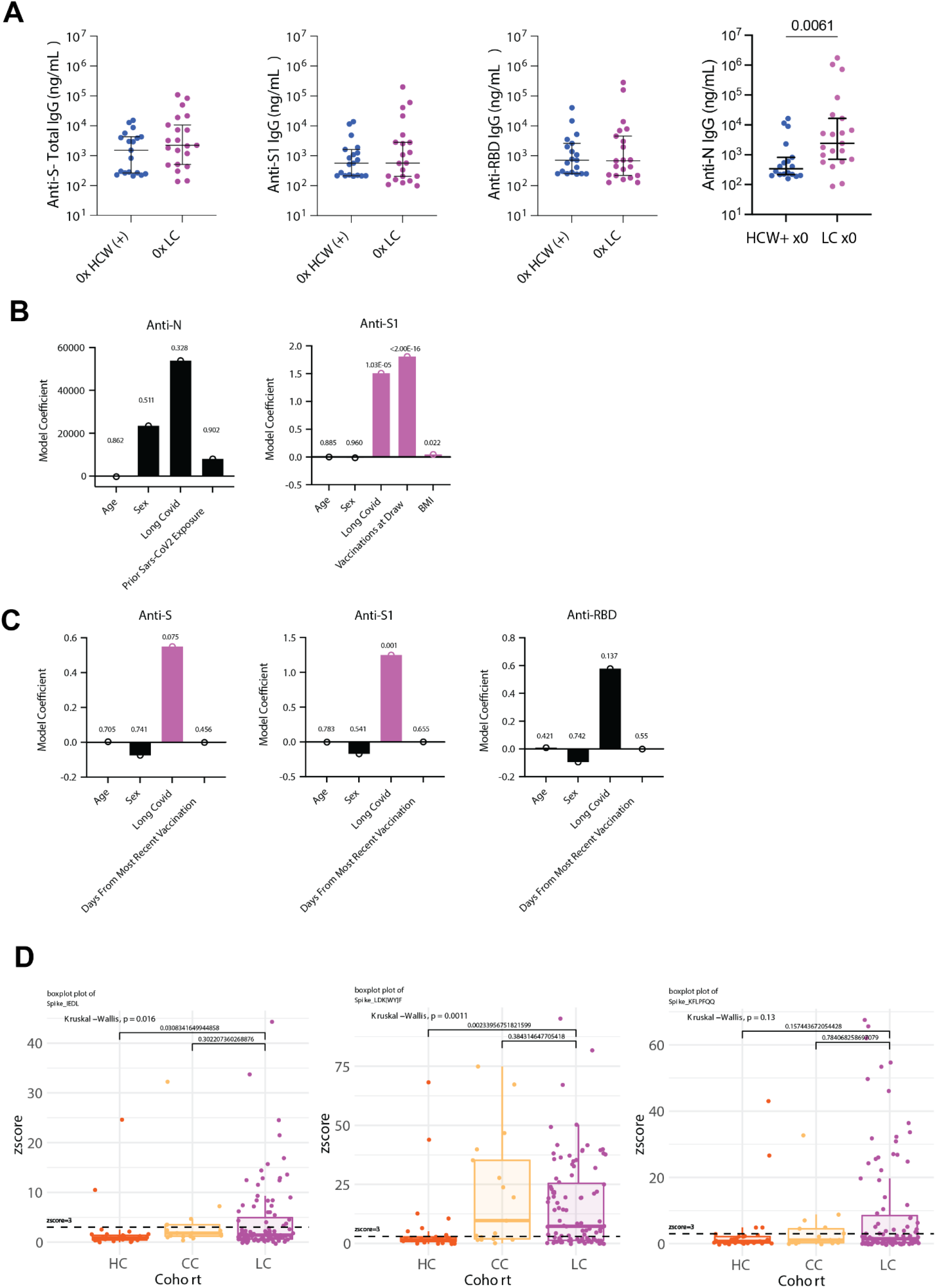
Analysis of SARS-CoV-2 specific antibodies. **(A)** Dot plots of IgG concentrations from historical, unvaccinated SARS-CoV-2 exposed controls (HCW+) and unvaccinated Long COVID participants. Vaccination status for each group is indicated by the form “x0” where the digit indicates the number of SARS-CoV-2 vaccine doses. Central bars indicate group means, and error bars represent 95% confidence interval estimates of group means. Significance for difference in group medians was assessed using Kruskal-Wallis with Dunn’s test for post-hoc comparison. Reported p-values are adjusted for multiple comparison using Tukey’s Method. **(B-C)** Results from generalized linear models of various SARS-CoV-2 antibody responses. Black bars are non-significant model predictors. Purple bars are significant model predictors. Model predictors are reported along the x-axis and included age, sex (categorical), Long COVID status (categorical), body mass index (BMI), prior SARS-CoV-2 exposure (anti-N only), and days from most recent vaccine (among vaccinated participants only). P-values are reported over individual predictor bars. **(D)** Box-plots of antibody binding to various SARS-CoV-2 linear peptide sequences. Central bars represent groups medians, with top and bottom bars representing 75^th^ and 25^th^ percentiles respectively. Significance for difference in group medians was assessed using Kruskal-Wallis with Dunn’s test for post-hoc comparison. Reported p-values are adjusted for multiple comparison using Tukey’s Method.

**Extended Figure S4.**
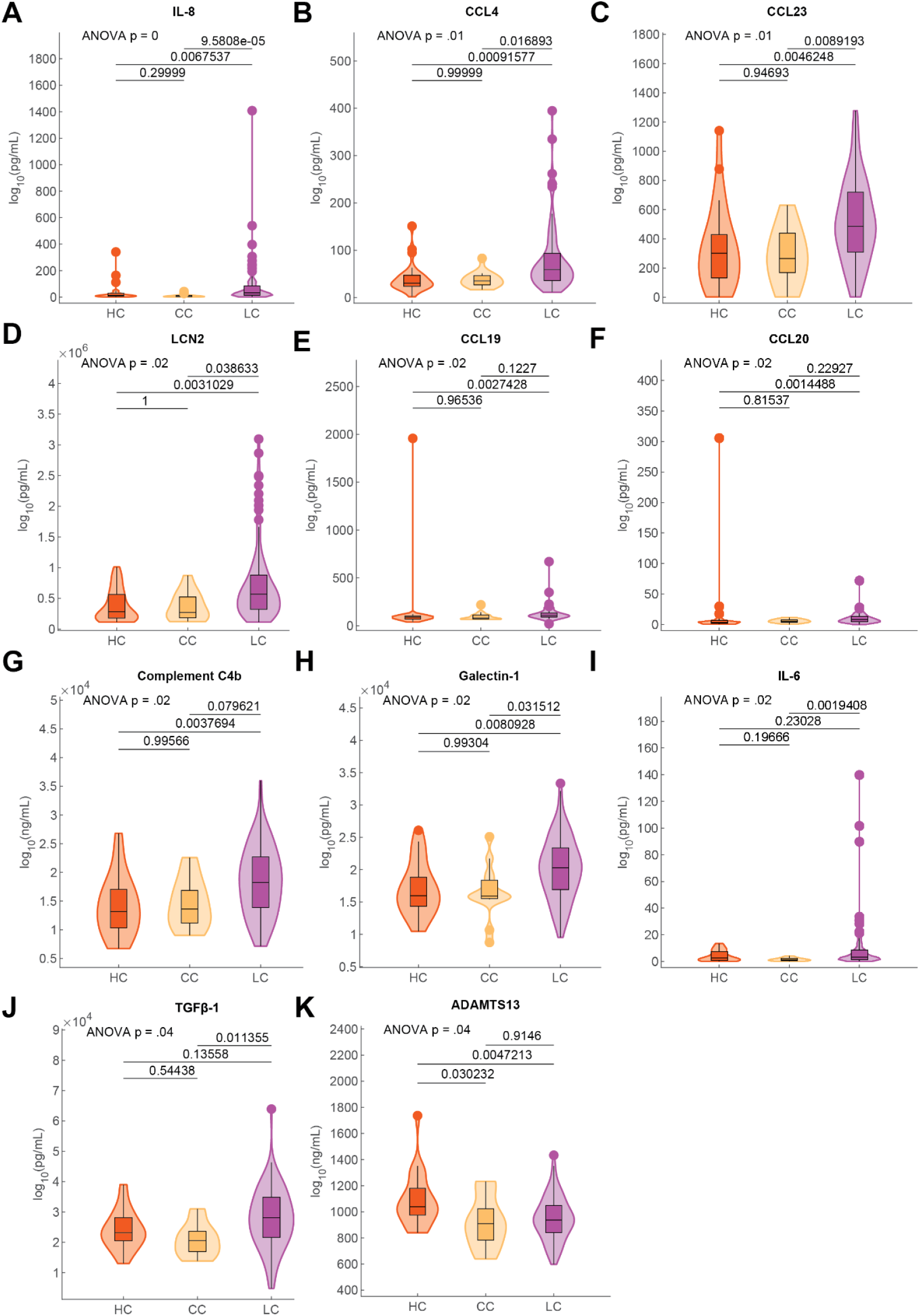
Significantly different soluble plasma factors across MY-LC groups. Violin plots of various circulating plasma factors across healthy (HC), convalescent (CC), and Long COVID (LC) groups. Significance for differences in group medians was assessed using Kruskal-Wallis with Dunn’s test for post-hoc comparisons. P-values were adjusted using Tukey’s method and shown where significant.

**Extended Figure S5.**
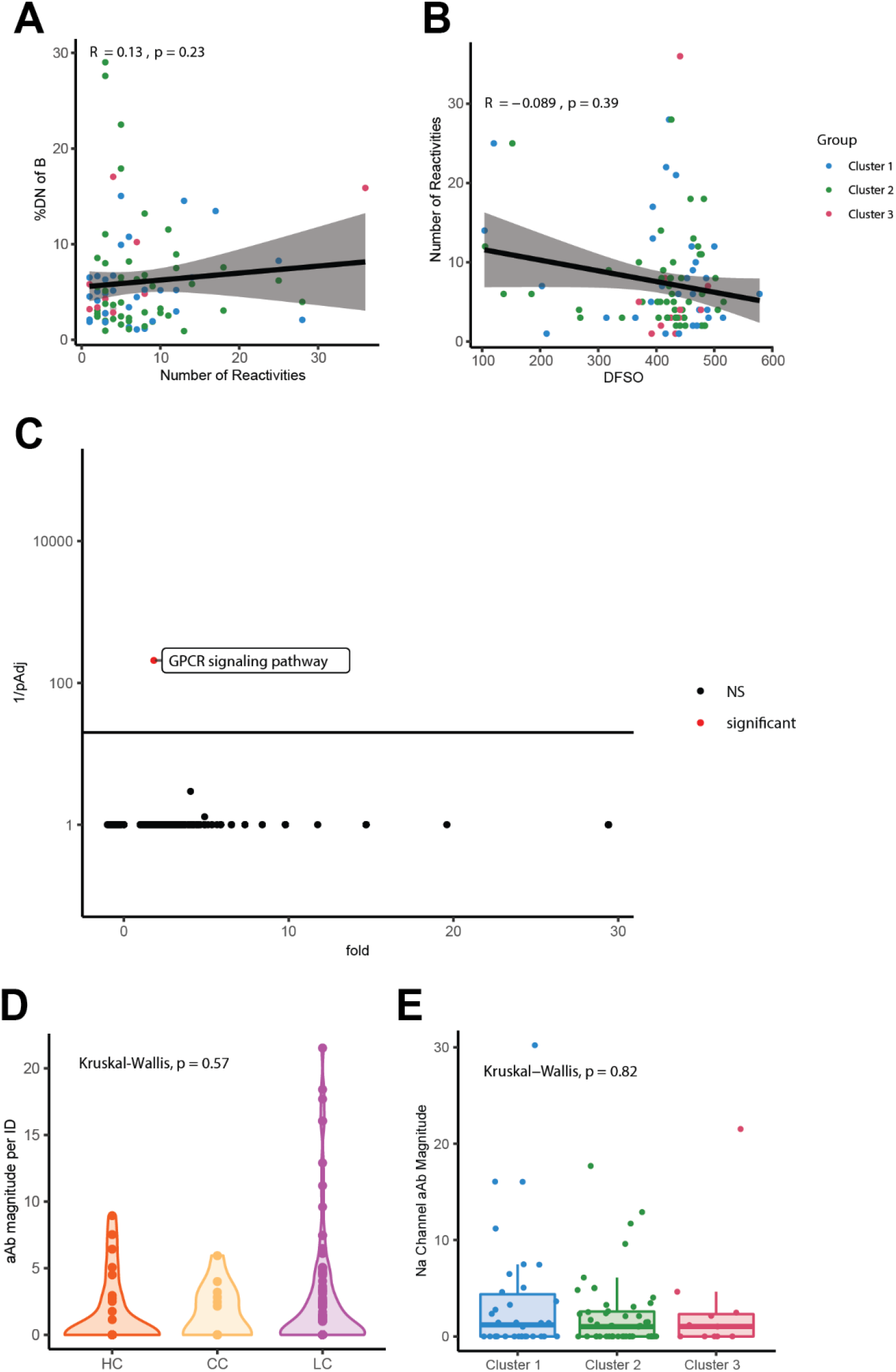
Analysis of private autoantibodies within the MY-LC Study. **(A,B)** Correlation plots depicting the relationship between number of autoantibody reactivities and % double negative (%DN) of B cells **(A)** or days from symptom onset (DFSO) and number of autoantibody reactivities **(B)**. For all correlations, Spearman’s Rho is reported with corresponding p-values. Black line depicts linear regression with 95% CI shaded. Colors depict Long COVID cluster (Cluster 3= blue; Cluster 2 = green; Cluster 1 = red). Each dot represents one individual. **(C)** Panther GO Biological Process over-representation analysis for reactivities unique to controls relative to the background REAP library proteins. Statistical significance determined by Fisher’s exact test with correction for multiple hypotheses by Bonferroni. Red color indicates adjusted p value <0.05. **(D)** REAP score reactivity magnitude against proteins belonging to the Go Process “sodium ion transport”. Reactivity magnitude is calculated as the sum of REAP scores for all reactivities per individual in a given Go Process domain. Statistical significance assessed by Kruskal-Wallis. Each dot represents one individual. **(E)** REAP score reactivity magnitude against proteins belonging to the GO Process “sodium ion transport” by LCPS cluster. Significance assessed by Kruskal-Wallis. Each dot represents one individual. Middle values of box-plot indicate group medians, with top and bottom lines depicting 75^th^ to 25^th^ percentiles, respectively.

**Figure S6.**
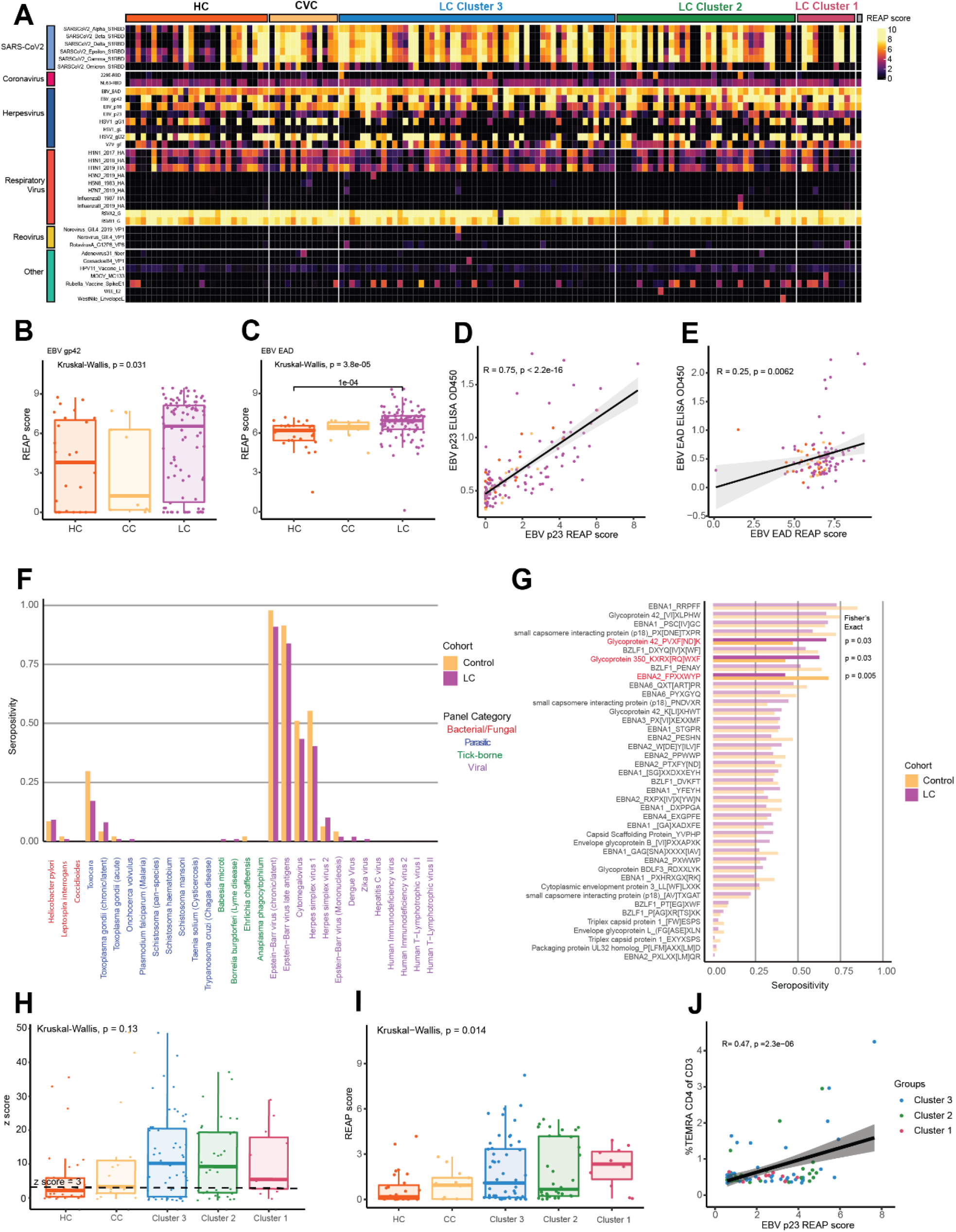
Non-SARS-CoV-2 humoral responses among participants with Long COVID. **(A)** Heatmap depicting REAP reactivities to viral antigens across the MY-LC study. Each column is one participant, with participants grouped by study group and LCPS. Column clustering within groups performed by K-means clustering. Each row is one viral protein. Reactivities depicted have at least one patient with a REAP score >= 1. **(B, C)** REAP scores for EBV gp42 **(B)** and EBV EAD **(C)** by group (HC = healthy control; CC = convalescent control; LC = Long Covid). Statistical significance determined by Kruskal Wallis. Post-hoc tests performed using Dunn’s test with Holm’s method to adjust for multiple comparisons. Each dot represents one individual. Box-plot depicts 25^th^ to 75^th^ percentile of the data, with the middle line representing the median. **(D, E)** Correlation plot depicting the relationship between EBV p23 **(D)** or EBV EAD **(E)** REAP score and EBV p23 or EBV EAD ELISA O.D. 450 nm. Statistical significance assessed by Pearson correlation. Black line depicts linear regression with 95% CI shaded. Colors depict group (pink = LC, yellow = CC, orange = HC). Each dot represents one individual. **(F)** Proportion of each group seropositive for each of 30 common pathogen panels as determined by SERA, grouped by pathogen-type (LC = Long COVID). Statistical significance determined by Fisher’s Exact test. **(G)** Proportion of each group seropositive (z-score ≥ 3) for each of 46 conserved linear motifs comprising the EBV disease panels. Motifs with significantly different seropositivity between groups are highlighted. Statistical significance determined by Fisher’s Exact test. **(H, I)** EBV gp42 PVXF[ND]K (S11H) or EBV p23 REAP score **(I)** grouped by participant group (HC = healthy control; CC = convalescent control; LC = Long Covid). Statistical significance determined by Kruskal Wallis. Post-hoc tests performed using Dunn’s test with Holm’s method to adjust for multiple comparisons. Each dot represents one individual. Boxplot colored box depicts 25^th^ to 75^th^ percentile of the data, with the middle line representing the median. **(J)** Correlation plot depicting the relationship between EBV p23 and %CD4+ T_EMRA_ of CD3+. Statistical significance assessed by Pearson correlation. Black line depicts linear regression with 95% CI shaded. Colors depict Long COVID Cluster (Cluster 1 = red; Cluster 2 = green; Cluster 3 = blue). Each dot represents one individual.

**Extended Figure S7.**
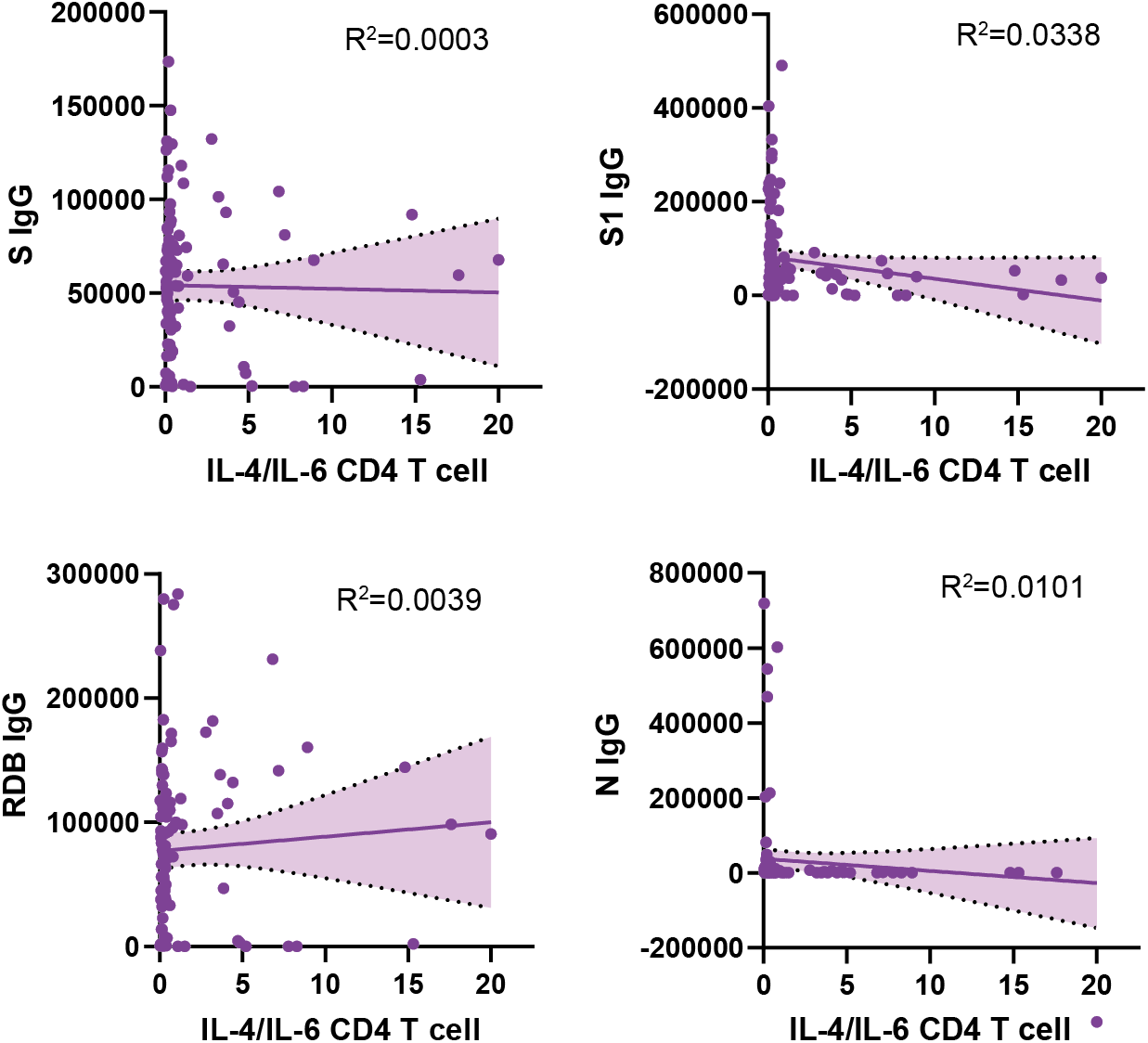
Lack of significant correlations of IL-4 / IL-6 double positive immune effectors with SARS-CoV-2 antibody responses. Correlation plots depicting the relationship between IL-4/IL-6 double positive T cells populations and various SARS-CoV-2 specific antibody responses. Black lines depict linear regression with 95% CI shaded.

**Extended Figure S8.**
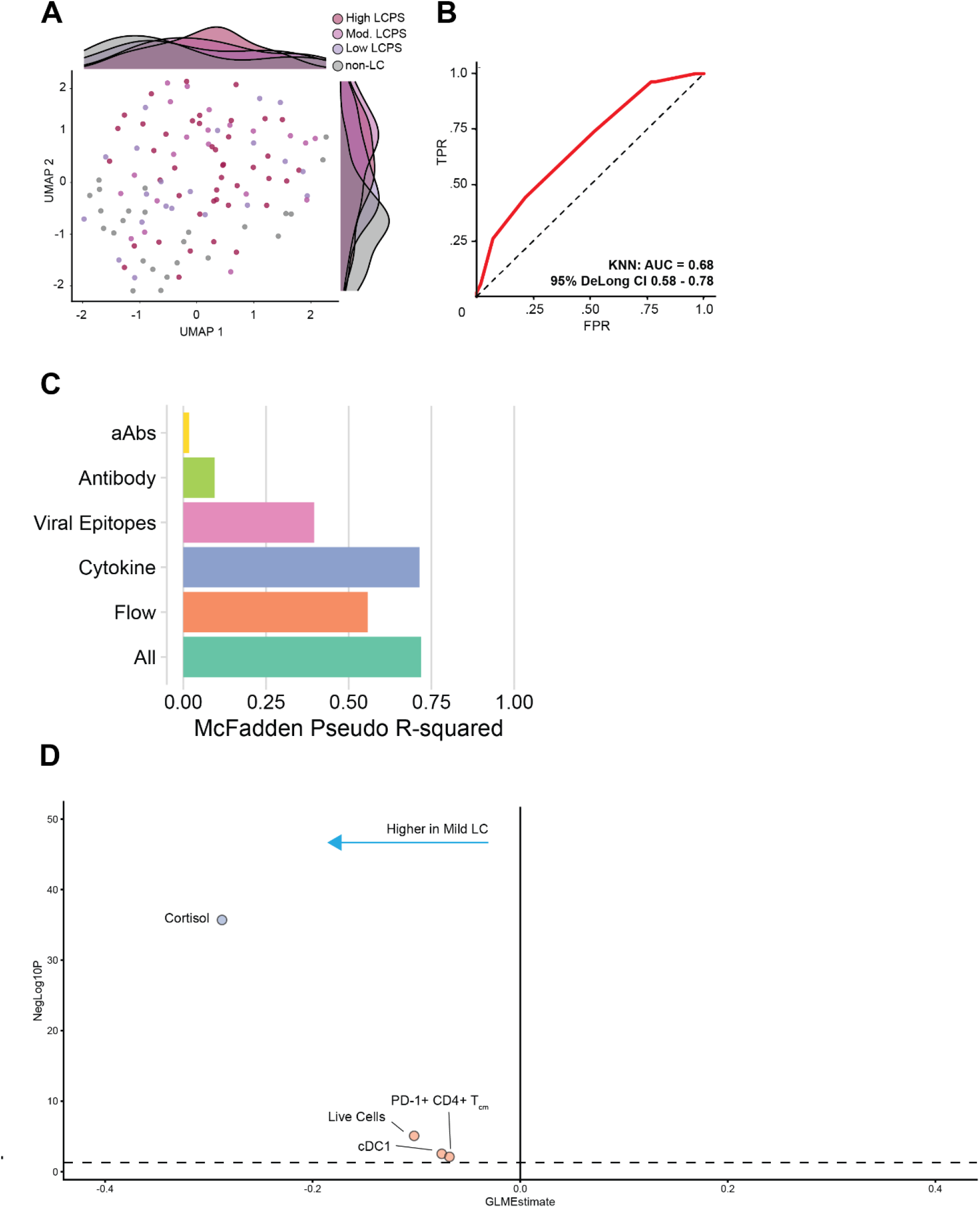
Principal components regression for prediction of LCPS scores. **(A)** UMAP projection of all collected immunological data of participants stratified by LCPS scores. **(B)** ROC curve analysis from unsupervised K-nearest neighbors (KNN) classification of individuals with high LCPS scores (> 80^th^ percentile). AUC and 95% CI intervals (DeLong’s Method) are reported. **(C)** McFadden’s pseudo R-squared are reported as bar plot for each data dimension. An integrated, parsimonious McFadden’s pseudo R-squared is reported for the final classification model (‘All’). **(D**) Dot-plot of immunologic features differentiating participants with mild LCPS scores from those with high LCPS score. Dots are colored according to individual data segments: orange = Flow cytometry, blue = Plasma cytokines, pink = viral epitopes, green = SARS-CoV-2 specific antibodies, yellow = autoantibodies to human exoproteome (aAbs).

**Extended Figure S9.**
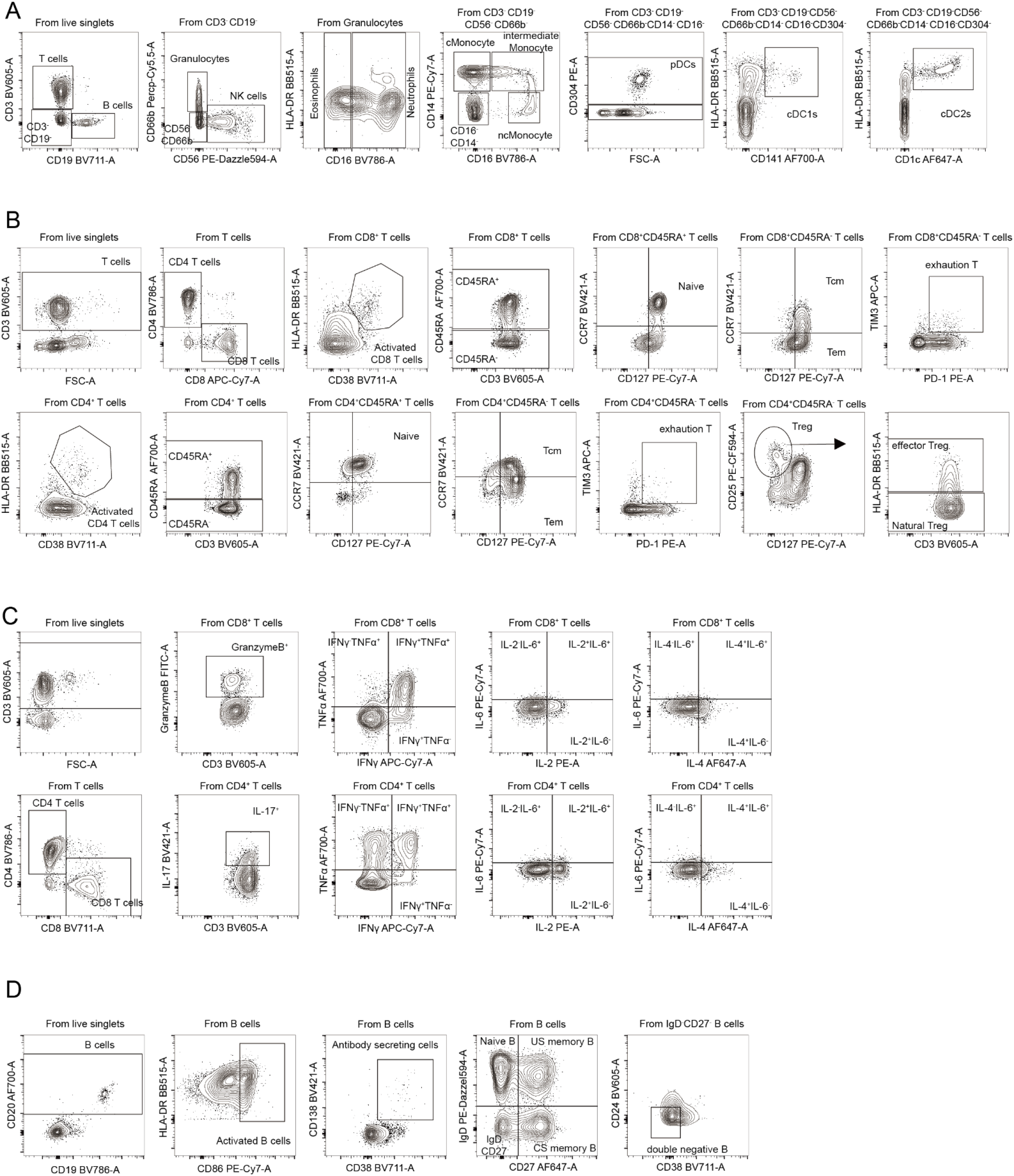
Flow Cytometry gating schematics. **(A-D).** Various gating strategies for granulocyte and myeloid populations **(A)**, T lymphocytes **(B)**, intracellular cytokine staining **(C),** and B lymphocytes **(D)**. A participant from the Long COVID group is shown as an example.

## Extended Tables

**Extended Table 1.**
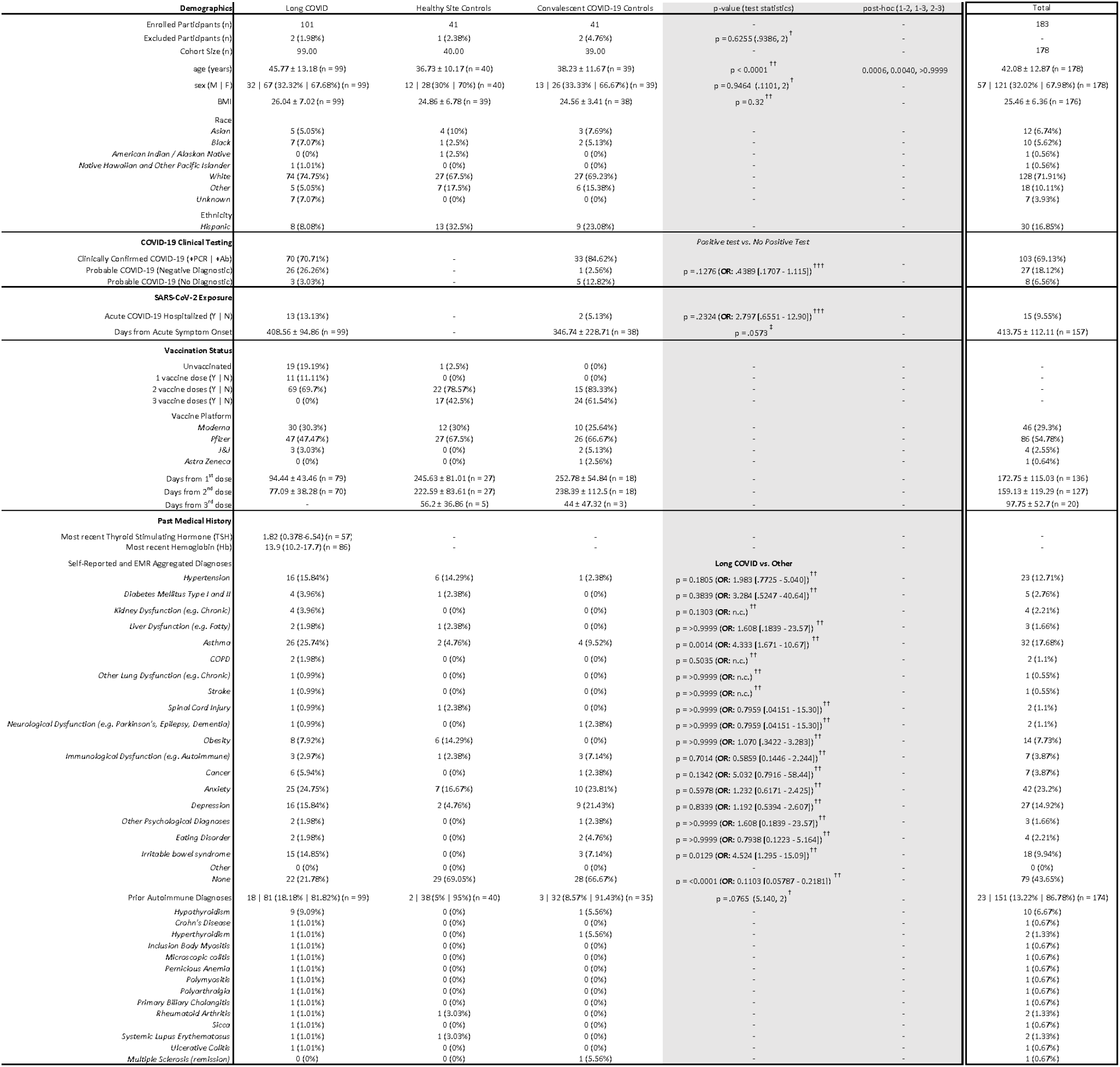
Clinical Demographics of MY-LC Study. Summary demographic and clinical characteristics for the MY-LC Study (Mount Sinai Study Groups). Participants were stratified into three study arms at enrollment: (1) Long COVID (prior SARS-CoV-2 infection with persistent, unexplained symptoms); (2) healthy study site control group (no prior SARS-CoV-2 infection); or (3) convalescent COVID-19 group (prior SARS-CoV-2 infection without persistent symptoms). Various demographic features and clinical characteristics are reported by row for each study group (row measurement units are specified in parentheses). Within each cell, participant counts or clinical feature averages are reported, with sample standard deviations, relative group percentages, and total numbers reported where pertinent. Results from statistical tests are reported as p-values and accompanying test statistics: † Chi-square test p-value (Chi-square test statistic, degrees of freedom (df)); †† Kruskal-Wallis ANOVA p-value; ††† Fisher’s Exact Test p-value (Odd’s Ratio: [95% Confidence Interval (Baptista-Pike)]); ‡ Mann-Whitney U test p-value. Post-hoc comparisons were conducted using Dunn’s test with Tukey’s correction for multiple comparison (column comparison order left-right: 1-2, 1-3, 2-3). Participant medical histories were collected and collated from binary self-reports of prior medical history and review of electronic medical records by study staff (positive responses in either participant self-report or EMR review were considered an overall binary positive response). Abbreviations: *n = number; M = male; F = female; BMI = body mass index; +PCR = positive result from SARS-CoV-2 nucleic acid test; +Ab = positive result from SARS-CoV-2 antibody test; Y= Yes; N = No*.

**Extended Table 2.** Normalized survey responses across MY-LC study groups. Survey responses for participants are organized by individual instruments (columns) and MY-LC groups (rows). Participant responses for each survey instrument were summed and normalized using standard min-max normalization procedures such that a value of 1 equals the maximum possible aggregate score and 0 equals the minimum possible aggregate score. Additionally, individual survey elements were oriented through inversion such that higher normalized scores on each instrument indicate a higher or worsened intensity for survey prompts. For each group, median values are displayed.

| Cohort | EuroQol EQ-5D-5L | EuroQol EQ-5D-5L VAS | MRC Breathlessness Scale | Fatigue Severity Scale | Fatigue VAS | PHQ-2 | GAD-7 | Pain VAS | Single-Item Sleep Quality Scale | Neuro-QOL v2.0 Cognitive Function Short Form | DSQ-PEM Short Form |
| --- | --- | --- | --- | --- | --- | --- | --- | --- | --- | --- | --- |
| HC | 0.00 | 0.10 | 0.00 | 0.15 | 0.12 | 0.00 | 0.10 | 0.00 | 0.30 | 0.14 | 0.00 |
| CC | 0.05 | 0.13 | 0.00 | 0.17 | 0.10 | 0.00 | 0.07 | 0.00 | 0.30 | 0.21 | 0.05 |
| LC | 0.30 | 0.35 | 0.25 | 0.83 | 0.69 | 0.17 | 0.29 | 0.50 | 0.50 | 0.43 | 0.50 |

**Extended Table 3.**
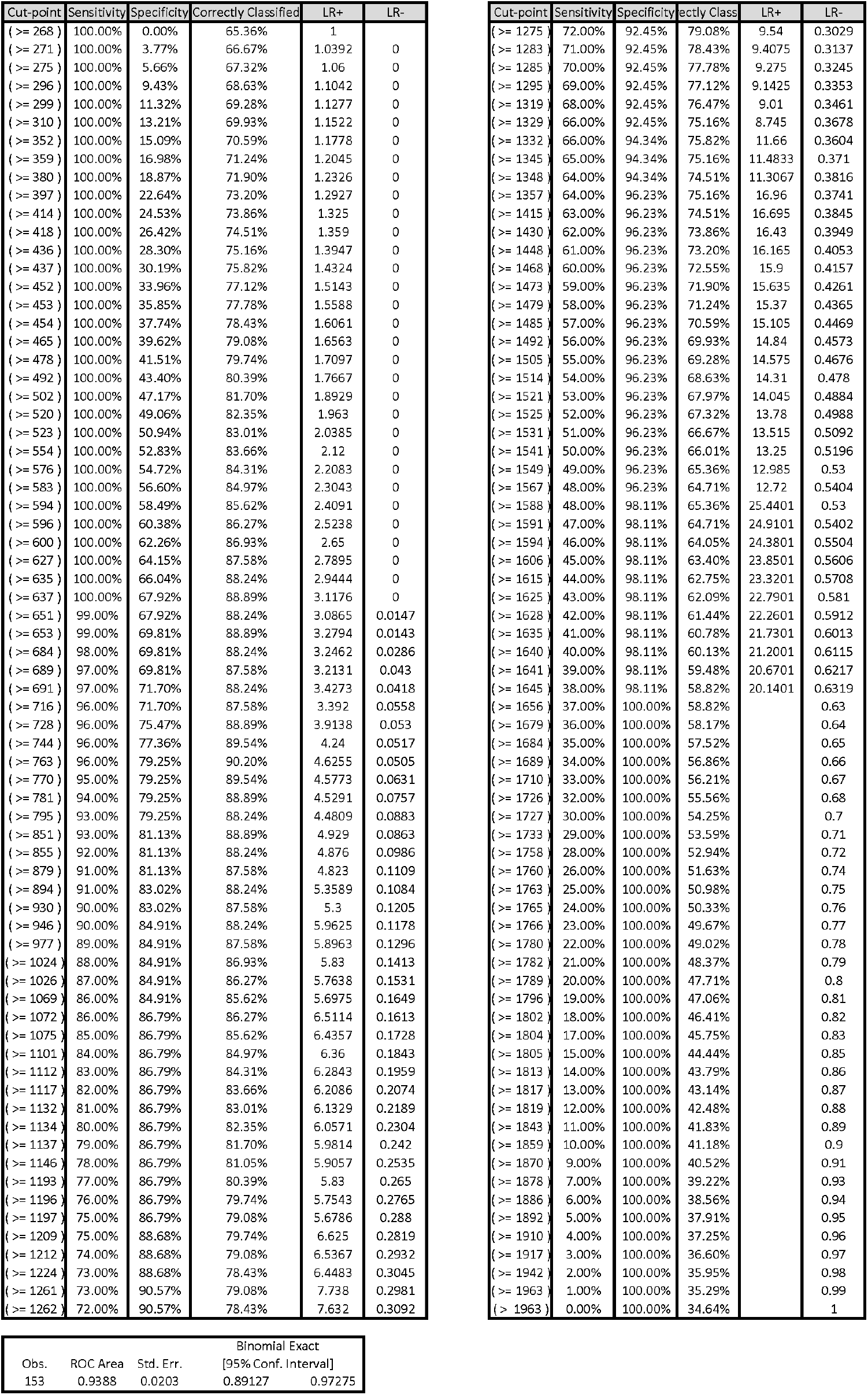
Determinations of optimal LCPS threshold. Classification metrics across different LCPS thresholds (‘Cut-offs’) (*Upper table*). Summary area-under the curve (AUC) statistics and bootstrap confidence intervals for Receiver-Operator curve analysis (ROC) (*lower table*)

**Extended Table 4.**
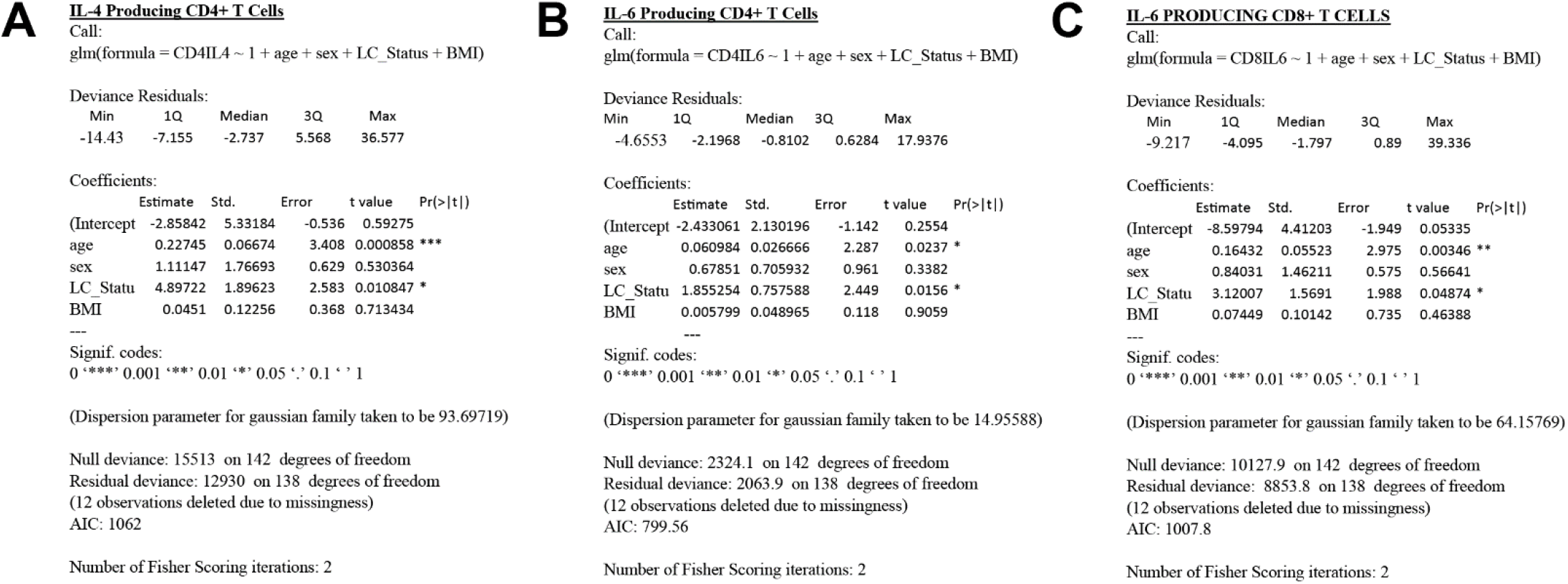
Statistical modeling of intracellular cytokine production. **(A-C)** Detailed generalized linear modeling results are reported for various cytokine producing T cell populations analyzed by flow cytometry.

**Extended Table 5.** Statistical modeling of anti-SARS-CoV-2 antibody responses. **(A-D)** Detailed generalized linear modeling results are reported for SARS-CoV-2 specific antibody responses with corresponding model formulations.

| Min | 1Q | Median | 3Q | Max |
| --- | --- | --- | --- | --- |
| -6.0196 | -0.3531 | 0.1937 | 0.6152 | 3.3648 |

Coefficients:
|  | Estimate | Std. Error | t value | Pr(>\|t\|) |
| --- | --- | --- | --- | --- |
| (Intercept) | 6.268002 | 0.84236 | 7.441 | 1.08E-11 *** |
| age | 0.002895 | 0.009257 | 0.313 | 0.755001 |
| sex | 0.037905 | 0.243218 | 0.156 | 0.876389 |
| LC_Status | 1.02332 | 0.281396 | 3.637 | 0.000393 *** |
| VAD | 1.343194 | 0.153473 | 8.752 | 7.88E-15 *** |
| BMI | 0.026281 | 0.017328 | 1.517 | 0.131713 |

| Min | 1Q | Median | 3Q | Max |
| --- | --- | --- | --- | --- |
| -6.0834 | -0.7336 | 0.0091 | 1.0018 | 4.0686 |

Coefficients:
|  | Estimate | Std. Error | t value | Pr(>\|t\|) |
| --- | --- | --- | --- | --- |
| (Intercept) | 4.511837 | 0.984926 | 4.581 | 1.05E-05 *** |
| age | 0.001579 | 0.010824 | 0.146 | 0.8842 |
| sex | -0.01428 | 0.284382 | -0.05 | 0.96 |
| LC_Status | 1.50799 | 0.329021 | 4.583 | 1.04E-05 *** |
| VAD | 1.810277 | 0.179447 | 10.088 | <2.0 E-16 *** |
| BMI | 0.047101 | 0.020261 | 2.325 | 0.0216 * |

| Min | 1Q | Median | 3Q | Max |
| --- | --- | --- | --- | --- |
| -5.9638 | -0.7555 | 0.152 | 0.8619 | 5.1572 |

Coefficients:
|  | Estimate | Std. Error | t value | Pr(>\|t\|) |
| --- | --- | --- | --- | --- |
| (Intercept) | 4.64593 | 1.01722 | 4.567 | 1.11E-05 *** |
| age | 0.0108 | 0.01118 | 0.966 | 0.33567 |
| sex | 0.08814 | 0.29371 | 0.3 | 0.76457 |
| LC_Status | 1.10777 | 0.33981 | 3.26 | 0.00141 ** |
| VAD | 1.87848 | 0.18533 | 10.136 | <2.0 E-16 *** |
| BMI | 0.03811 | 0.02093 | 1.821 | 0.07082 |

| Min | 1Q | Median | 3Q | Max |
| --- | --- | --- | --- | --- |
| -3.5985 | -1.1907 | -0.2953 | 0.7132 | 6.026 |

Coefficients:
|  | Estimate | Std. Error | t value | Pr(>\|t\|) |
| --- | --- | --- | --- | --- |
| (Intercept) | 4.57123 | 1.226254 | 3.728 | 0.000284 *** |
| age | 0.003411 | 0.013476 | 0.253 | 0.800565 |
| sex | 0.117323 | 0.354061 | 0.331 | 0.740888 |
| LC_Status | 0.166987 | 0.409638 | 0.408 | 0.684185 |
| VAD | -0.50522 | 0.223416 | -2.261 | 0.025348 * |
| BMI | 0.113647 | 0.025225 | 4.505 | 1.43E-05 *** |

Signif. codes: 0 '\*\*\*' 0.001 '\*\*' 0.01 '\*' 0.05 '.' 0.1 ' ' 1
(Dispersion parameter for gaussian family taken to be 3.761342)
Null deviance: 624.13 on 139 degrees of freedom Residual deviance: 504.02 on 134 degrees of freedom (15 observations deleted due to missingness) AIC: 590.64
Number of Fisher Scoring iterations: 2

**Extended Table 6.**
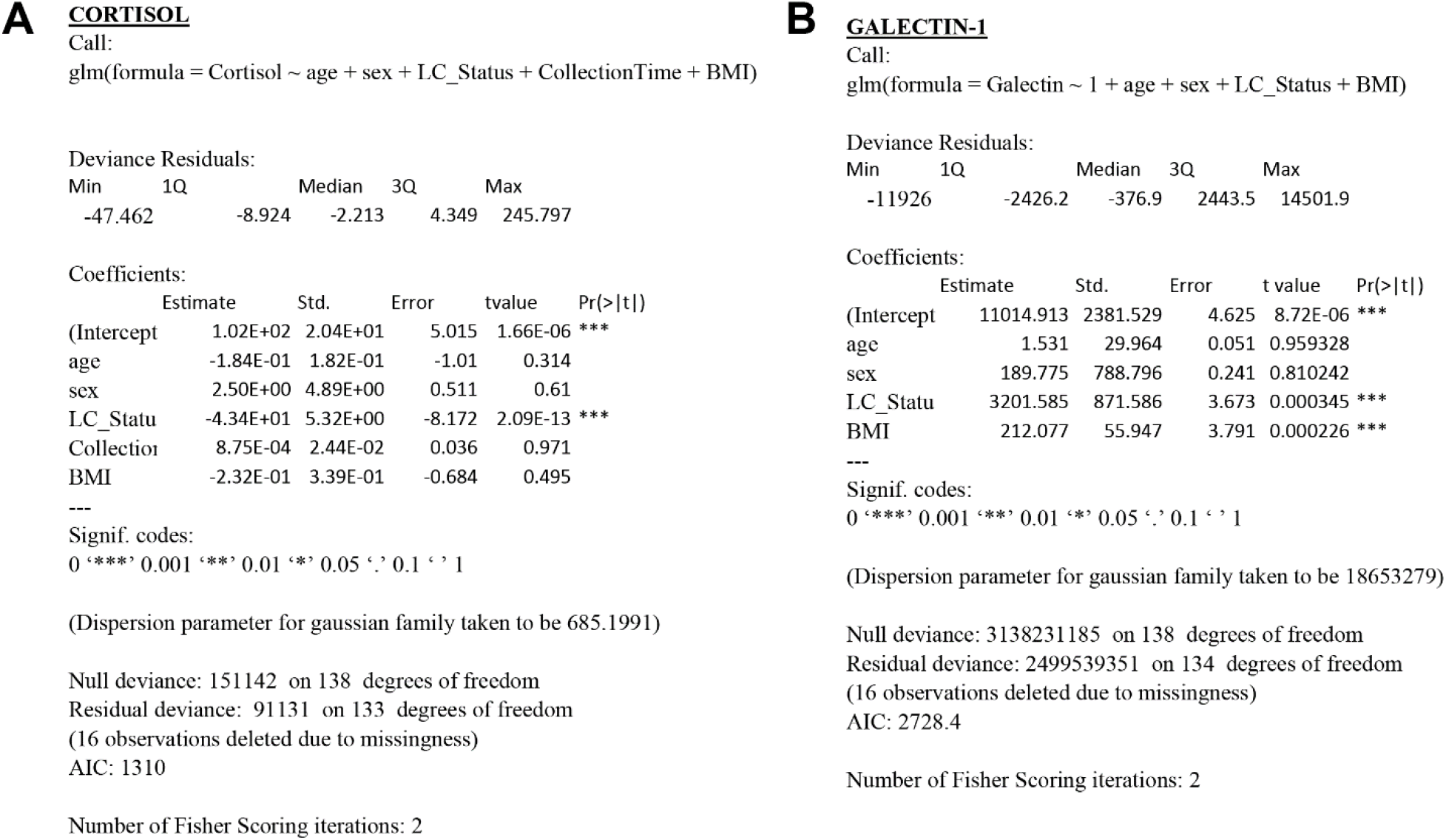
Statistical modeling of circulating plasma factors. **(A,B)** Detailed generalized linear modeling results are reported for soluble plasma factors.

**Extended Table 7.**
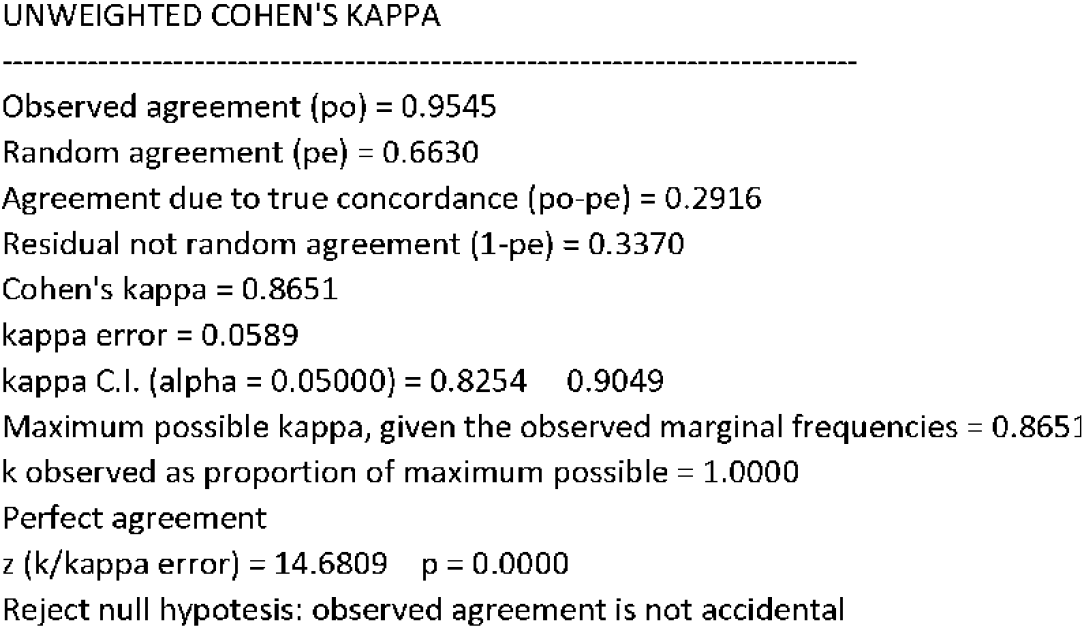
Cohen’s Kappa calculation for LCPS and Machine Learning classifications of Long COVID status.

## Supplementary Information

**Supplementary Table 1.**
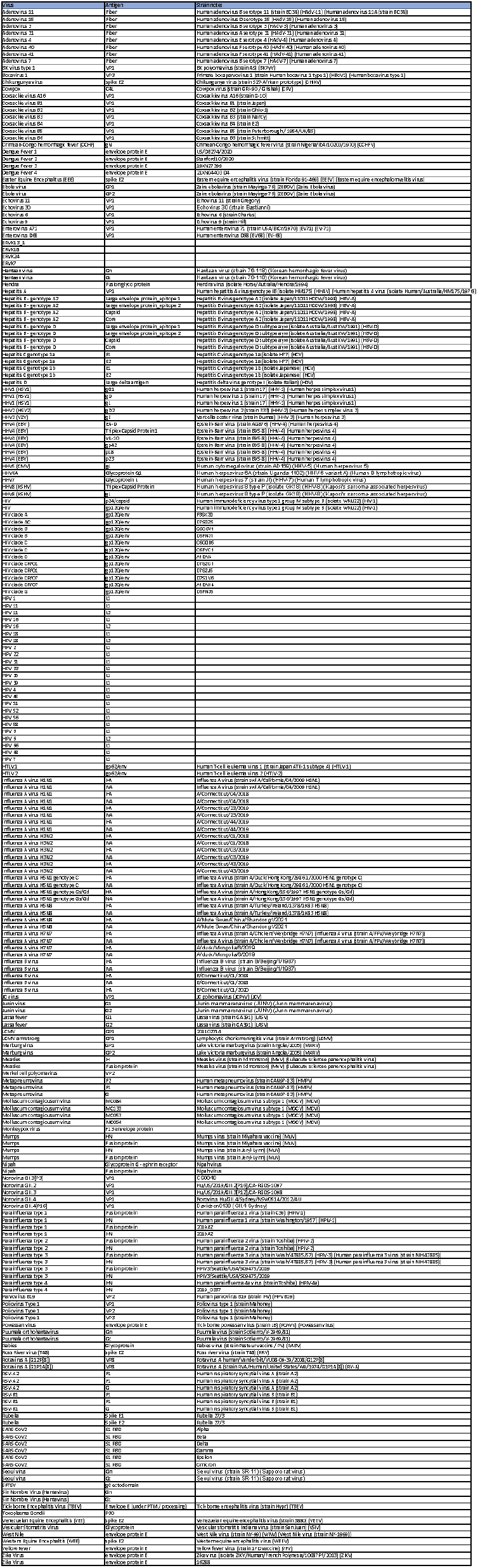
Viral antigens included in REAP analysis.

**Supplementary Table 2.**
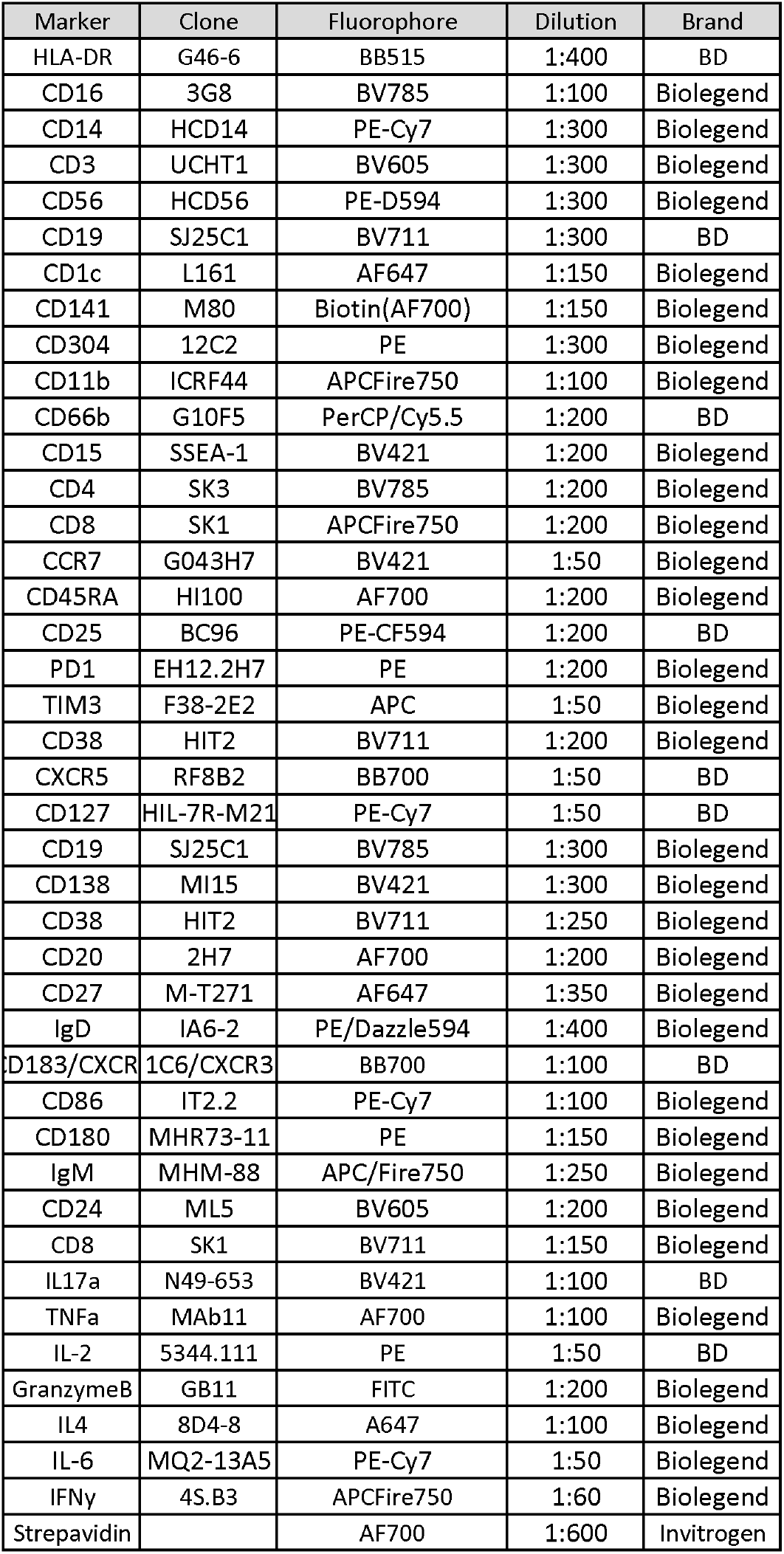
Antibody clones and dilutions used for flow cytometry analysis of PBMC populations.

## References

1. Michelen, M., et al. Characterising long COVID: A living systematic review. BMJ Global Health vol. 6 Preprint at https://doi.org/10.1136/bmjgh-2021-005427 (2021).

2. Al-Aly, Z., Xie, Y. & Bowe, B. High-dimensional characterization of post-acute sequelae of COVID-19. Nature 594, 259–264 (2021).

3. Davis, H. E., et al. Characterizing long COVID in an international cohort: 7 months of symptoms and their impact. eClinicalMedicine 38, (2021).

4. Clark, D. V., et al. Long-term sequelae after Ebola virus disease in Bundibugyo, Uganda: A retrospective cohort study. The Lancet Infectious Diseases 15, 905–912 (2015).

5. Kalifa Diallo, M. S., et al. Understanding Long-term Evolution and Predictors of Sequelae of Ebola Virus Disease Survivors in Guinea: A 48-month prospective, longitudinal cohort study (PosteboguI). Clinical Infectious Diseases 73, 2166–2174 (2021).

6. Wiedemann, A. et al. Long-lasting severe immune dysfunction in Ebola virus disease survivors. Nature Communications 11, 1–11 (2020).

7. Rowe, A. K. et al. Clinical, virologic, and immunologic follow-up of convalescent Ebola hemorrhagic fever patients and their household contacts, Kikwit, Democratic Republic of the Congo. Journal of Infectious Diseases 179, (1999).

8. García, G. et al. Long-term persistence of clinical symptoms in dengue-infected persons and its association with immunological disorders. International Journal of Infectious Diseases 15, 38–43 (2011).

9. Patel, H., Sander, B. & Nelder, M. P. Long-term sequelae of West Nile virus-related illness: A systematic review. The Lancet Infectious Diseases 15, 951–959 (2015).

10. Paixão, E. S. et al. Chikungunya chronic disease: a systematic review and meta-analysis. Transactions of The Royal Society of Tropical Medicine and Hygiene 112, 301–316 (2018).

11. Sejvar, J. J. et al. Long-term neurological and functional outcome in Nipah virus infection. Annals of Neurology 62, 235–242 (2007).

12. Trojan, D. A. & Cashman, N. R. Post-poliomyelitis syndrome. Muscle and Nerve 31, 6–19 (2005).

13. Hickie, I. et al. Post-infective and chronic fatigue syndromes precipitated by viral and non-viral pathogens: Prospective cohort study. British Medical Journal 333, 575–578 (2006).

14. Gowers, W. R. A Post-Graduate Lecture ON THE NERVOUS SEQUELAE OF INFLUENZA. The Lancet 142, 73–76 (1893).

15. Sisley, R. INFLUENZA: A NOTE ON ITS TREATMENT AND ON THAT OF ITS SEQUELAE. The Lancet 135, 167 (1890).

16. Nicholson, F. The Complications and Sequelae of Influenza. BMJ 1, 1273–1275 (1891).

17. Althaus, J. Influenza: Its Pathology, Symptoms, Complications, and Sequels: Its Origin and Mode of Spreading; and Its Diagnosis, Prognosis, and Treatment. (Longmans, 1892).

18. Choutka, J., Jansari, V., Hornig, M. & Iwasaki, A. Unexplained post-acute infection syndromes. Nature Medicine 28, 911–923 (2022).

19. Dong, E., Du, H. & Gardner, L. An interactive web-based dashboard to track COVID-19 in real time. The Lancet Infectious Diseases vol. 20 533–534 Preprint at https://doi.org/10.1016/S1473-3099(20)30120-1 (2020).

20. Lucas, C. et al. Delayed production of neutralizing antibodies correlates with fatal COVID-19. Nature Medicine 27, 1178–1186 (2021).

21. Wang, E. Y. et al. Diverse functional autoantibodies in patients with COVID-19. Nature 595, 283– 288 (2021).

22. Lucas, C. et al. Longitudinal analyses reveal immunological misfiring in severe COVID-19. Nature 584, 463–469 (2020).

23. Mathew, D. et al. Deep immune profiling of COVID-19 patients reveals distinct immunotypes with therapeutic implications. Science (1979) 369, (2020).

24. Gupta, A. et al. Extrapulmonary manifestations of COVID-19. Nature Medicine vol. 26 1017–1032 Preprint at https://doi.org/10.1038/s41591-020-0968-3 (2020).

25. Xie, Y., Xu, E., Bowe, B. & Al-Aly, Z. Long-term cardiovascular outcomes of COVID-19. Nature Medicine 28, 583–590 (2022).

26. Nalbandian, A. et al. Post-acute COVID-19 syndrome. Nature Medicine 27, 601–615 (2021).

27. Logue, J. K. et al. Sequelae in Adults at 6 Months after COVID-19 Infection. JAMA Network Open 4, (2021).

28. Huang, C. et al. 6-month consequences of COVID-19 in patients discharged from hospital: a cohort study. The Lancet 397, 220–232 (2021).

29. Tabacof, L. et al. Post-acute COVID-19 Syndrome Negatively Impacts Physical Function, Cognitive Function, Health-Related Quality of Life, and Participation. Am J Phys Med Rehabil 101, 48–52 (2022).

30. Groff, D. et al. Short-term and Long-term Rates of Postacute Sequelae of SARS-CoV-2 Infection: A Systematic Review. JAMA Network Open 4, (2021).

31. Chen, C. et al. Global Prevalence of Post COVID-19 Condition or Long COVID: A Meta-Analysis and Systematic Review. J Infect Dis (2022) doi:10.1093/infdis/jiac136.

32. Fernández-Castañeda, A. et al. Mild respiratory COVID can cause multi-lineage neural cell and myelin dysregulation. Cell (2022) doi:10.1016/j.cell.2022.06.008.

33. Gaebler, C. et al. Evolution of antibody immunity to SARS-CoV-2. Nature 591, 639–644 (2021).

34. Chertow, D. et al. SARS-CoV-2 infection and persistence throughout the human body and brain. doi:10.21203/rs.3.rs-1139035/v1.

35. Cheung, C. C. L. et al. Residual SARS-CoV-2 viral antigens detected in GI and hepatic tissues from five recovered patients with COVID-19. Gut 71, 226–229 (2022).

36. Arostegui, D., et al. Persistent SARS-CoV-2 Nucleocapsid Protein Presence in the Intestinal Epithelium of a Pediatric Patient 3 Months After Acute Infection. JPGN Reports 3, e152 (2022).

37. Su, Y. et al. Multiple early factors anticipate post-acute COVID-19 sequelae. Cell 185, 881–895.e20 (2022).

38. Mehandru, S. & Merad, M. Pathological sequelae of long-haul COVID. Nature Immunology vol. 23 194–202 Preprint at https://doi.org/10.1038/s41590-021-01104-y (2022).

39. Guilliams, M., Mildner, A. & Yona, S. Developmental and Functional Heterogeneity of Monocytes. Immunity vol. 49 595–613 Preprint at https://doi.org/10.1016/j.immuni.2018.10.005 (2018).

40. Wolf, A. A., Yáñez, A., Barman, P. K. & Goodridge, H. S. The ontogeny of monocyte subsets. Frontiers in Immunology vol. 10 Preprint at https://doi.org/10.3389/fimmu.2019.01642 (2019).

41. Narasimhan, P. B., Marcovecchio, P., Hamers, A. A. J. & Hedrick, C. C. Nonclassical Monocytes in Health and Disease. Annual Review of Immunology 37, 439–456 (2019).

42. Soto, J. A., et al. The Role of Dendritic Cells During Infections Caused by Highly Prevalent Viruses. Frontiers in Immunology vol. 11 Preprint at https://doi.org/10.3389/fimmu.2020.01513 (2020).

43. Ng, S. L., Teo, Y. J., Setiagani, Y. A., Karjalainen, K. & Ruedl, C. Type 1 Conventional CD103+ Dendritic Cells Control Effector CD8+ T Cell Migration, Survival, and Memory Responses During Influenza Infection. Frontiers in Immunology 9, (2018).

44. Ershler, W. B. & Keller, E. T. Age-Associated Increased Interleukin-6 Gene Expression, Late-Life Diseases, and Frailty. Annual Review of Medicine 51, 245–270 (2000).

45. Gardner, E. M. & Murasko, D. M. Age-related changes in Type 1 and Type 2 cytokine production in humans. Biogerontology 3, 271–290 (2002).

46. Pieren, D. K. J., Smits, N. A. M., van de Garde, M. D. B. & Guichelaar, T. Response kinetics reveal novel features of ageing in murine T cells. Scientific Reports 9, (2019).

47. Li, Y. et al. Linear epitopes of SARS-CoV-2 spike protein elicit neutralizing antibodies in COVID-19 patients. Cellular and Molecular Immunology vol. 17 1095–1097 Preprint at https://doi.org/10.1038/s41423-020-00523-5 (2020).

48. Poh, C. M. et al. Two linear epitopes on the SARS-CoV-2 spike protein that elicit neutralising antibodies in COVID-19 patients. Nature Communications 11, (2020).

49. Wang, E. Y., et al. High-throughput identification of autoantibodies that target the human exoproteome. Cell Reports Methods 2, (2022).

50. Gold, J. E., Okyay, R. A., Licht, W. E. & Hurley, D. J. Investigation of long covid prevalence and its relationship to epstein-barr virus reactivation. Pathogens 10, (2021).

51. Kamath, K., et al. Antibody epitope repertoire analysis enables rapid antigen discovery and multiplex serology. Scientific Reports 10, (2020).

52. Metcalf, T. U. et al. Human Monocyte Subsets Are Transcriptionally and Functionally Altered in Aging in Response to Pattern Recognition Receptor Agonists. The Journal of Immunology 199, 1405–1417 (2017).

53. Swank, Z., Senussi, Y., Alter, G. & Walt, D. R. Persistent circulating SARS-CoV-2 spike is associated with post-acute COVID-19 sequelae. doi:10.1101/2022.06.14.22276401.

54. Charmandari, E., Nicolaides, N. C. & Chrousos, G. P. Adrenal insufficiency. The Lancet 383, 2152– 2167 (2014).

55. McKenzie, R. et al. Low-Dose Hydrocortisone for Treatment of Chronic Fatigue SyndromeA Randomized Controlled Trial. JAMA 280, 1061–1066 (1998).

56. Meng, M. et al. COVID-19 associated EBV reactivation and effects of ganciclovir treatment. Immunity, Inflammation and Disease 10, (2022).

57. Lanz, T. v, et al. Clonally expanded B cells in multiple sclerosis bind EBV EBNA1 and GlialCAM. Nature 603, 321–327 (2022).

58. Bjornevik, K., et al. MULTIPLE SCLEROSIS Longitudinal analysis reveals high prevalence of Epstein-Barr virus associated with multiple sclerosis. Science vol. 375 https://www.science.org (2022).

## Methods References

59. World Health Organization. Public health surveillance for COVID-19 Interim guidance Key points. WHO/2019-nCoV/SurveillanceGuidance/2022.1 (2022).

60. Krupp, L. B., LaRocca, N. G., Muir-Nash, J. & Steinberg, A. D. The Fatigue Severity Scale: Application to Patients With Multiple Sclerosis and Systemic Lupus Erythematosus. Archives of Neurology 46, 1121–1123 (1989).

61. Cotler, J., Holtzman, C., Dudun, C. & Jason, L. A Brief Questionnaire to Assess Post-Exertional Malaise. Diagnostics 8, 66 (2018).

62. Stenton, C. The MRC breathlessness scale. Occup Med (Lond) 58, 226–227 (2008).

63. Iverson, G. L., Connors, E. J., Marsh, J. & Terry, D. P. Examining normative reference values and item-level symptom endorsement for the quality of life in neurological disorders (Neuro-QoL^TM^) v2.0 Cognitive Function-Short Form. Archives of Clinical Neuropsychology 36, 126–134 (2021).

64. Herdman, M. et al. Development and preliminary testing of the new five-level version of EQ-5D (EQ-5D-5L). Quality of Life Research 20, 1727–1736 (2011).

65. Spitzer, R. L., Kroenke, K., Williams, J. B. W. & Löwe, B. A Brief Measure for Assessing Generalized Anxiety Disorder: The GAD-7. Archives of Internal Medicine 166, 1092–1097 (2006).

66. Kroenke, K., Spitzer, R. L. & Williams, J. B. W. The Patient Health Questionnaire-2: Validity of a Two-Item Depression Screener. Medical Care 41, (2003).

67. Snyder, E., Cai, B., DeMuro, C., Morrison, M. F. & Ball, W. A new single-item sleep quality scale: Results of psychometric evaluation in patients with chronic primary insomnia and depression. Journal of Clinical Sleep Medicine 14, 1849–1857 (2018).

68. Harris, P. A., et al. The REDCap consortium: Building an international community of software platform partners. Journal of Biomedical Informatics vol. 95 Preprint at https://doi.org/10.1016/j.jbi.2019.103208 (2019).

69. Harris, P. A. et al. Research electronic data capture (REDCap)-A metadata-driven methodology and workflow process for providing translational research informatics support. Journal of Biomedical Informatics 42, 377–381 (2009).

70. Haynes, W. A., Kamath, K., Waitz, R., Daugherty, P. S. & Shon, J. C. Protein-Based Immunome Wide Association Studies (PIWAS) for the Discovery of Significant Disease-Associated Antigens. Frontiers in Immunology 12, (2021).

71. Pantazes, R. J., et al. Identification of disease-specific motifs in the antibody specificity repertoire via next-generation sequencing. Scientific Reports 6, (2016).

72. Robinson, M. D., McCarthy, D. J. & Smyth, G. K. edgeR: A Bioconductor package for differential expression analysis of digital gene expression data. Bioinformatics 26, 139–140 (2009).

73. R Core Team. R: A language and environment for statistical computing. Preprint at (2018).

74. Wickham, H. ggplot2. (Springer New York, 2009). doi:10.1007/978-0-387-98141-3.

75. Becht, E. et al. Dimensionality reduction for visualizing single-cell data using UMAP. Nature Biotechnology 37, 38–44 (2019).

76. Robin, X. et al. pROC: An open-source package for R and S+ to analyze and compare ROC curves. BMC Bioinformatics 12, (2011).

77. DeLong, E. R., DeLong, D. M. & Clarke-Pearson, D. L. Comparing the Areas under Two or More Correlated Receiver Operating Characteristic Curves: A Nonparametric Approach. Biometrics 44, 837 (1988).

78. Gu, Z., Eils, R. & Schlesner, M. Complex heatmaps reveal patterns and correlations in multidimensional genomic data. Bioinformatics 32, 2847–2849 (2016).

79. Hennig, C. Flexible Procedures for Clustering. Preprint at https://cran.r-project.org/web/packages/fpc/index.html (2020).

